# BCG vaccination of healthcare workers does not reduce SARS-CoV-2 infections nor infection severity or duration: a randomised placebo-controlled trial

**DOI:** 10.1101/2022.12.12.22283282

**Authors:** Juana Claus, Thijs ten Doesschate, Cheyenne Gumbs, Cornelis H. van Werkhoven, Thomas W. van der Vaart, Axel B. Janssen, Gaby Smits, Rob van Binnendijk, Fiona van der Klis, Debbie van Baarle, Fernanda L. Paganelli, Helen Leavis, Lilly M. Verhagen, Simone A. Joosten, Marc J.M. Bonten, Mihai G. Netea, Janneke H. H. M. van de Wijgert, the BCG-Corona study group

**Author notes:** **Correspondence:** Janneke van de Wijgert, Julius Center for Health Sciences and Primary Care, University Medical Center Utrecht Universiteitsweg 100, Stratenum room 7.127, 3584 CG Utrecht.

## Abstract

**Background:** Bacillus Calmette-Guerin (BCG) vaccination has been hypothesised to reduce SARS-CoV-2 infection, severity, and/or duration via trained immunity induction.

**Methods:** Healthcare workers (HCWs) in 9 Dutch hospitals were randomised to BCG or placebo vaccination (1:1) in March/April 2020 and followed for one year. They reported daily symptoms, SARS-CoV-2 test results, and healthcare-seeking behaviour via a smartphone application, and donated blood for SARS-CoV-2 serology at two time points.

**Results:** 1,511 HCWs were randomised and 1,309 analysed (665 BCG and 644 placebo). Of the 298 infections detected during the trial, 74 were detected by serology only. The SARS-CoV-2 incidence rates were 0.25 and 0.26 per person-year in the BCG and placebo groups, respectively (incidence rate ratio=0.95; 95% confidence interval 0.76-1.21; p=0.732). Only three participants required hospitalisation for COVID-19. The proportions of participants with asymptomatic, mild, or mild-to-moderate infections, and the mean infection durations, did not differ between randomisation groups. Unadjusted and adjusted logistic regression and Cox proportional hazards models showed no differences between BCG and placebo vaccination for any of these outcomes either. The percentage of participants with seroconversion (7.8% versus 2.8%; p=0.006) and mean anti-S1 antibody concentration (13.1 versus 4.3 IU/ml; p=0.023) were higher in the BCG than placebo group at 3 months but not at 6 or 12 months post-vaccination.

**Conclusions:** BCG vaccination of HCWs did not reduce SARS-CoV-2 infections nor infection duration or severity (on a scale from asymptomatic to moderate). In the first 3 months after vaccination, BCG vaccination may enhance SARS-CoV-2 antibody production during SARS-CoV-2 infection.

## Introduction

The Bacillus Calmette-Guerin (BCG) vaccine is a widely used live-attenuated vaccine against tuberculosis. Routine childhood and/or healthcare worker (HCW) vaccination is performed worldwide, except in some countries with a low tuberculosis burden including the Netherlands. New interest in BCG has arisen after recent studies showed that BCG vaccination also has non-specific protective effects against other respiratory tract infections due to epigenetic and metabolic reprogramming of innate immune cells.^1,2^ This process is termed ‘trained immunity’. It was first observed in children in high infection prevalence settings,^3^ but later also in observational studies in adults,^4,5^ and in adults challenged with malaria or live-attenuated yellow fever or influenza vaccination after BCG vaccination.^6–8^

In the first year of the SARS-CoV-2 pandemic, testing and contact-tracing capacity, as well as personal protection equipment supplies, were limited and SARS-CoV-2 vaccines were not yet available. This raised challenges, especially for HCWs and vulnerable adults. HCWs were burdened with a high risk of exposure and infection while the demand for medical personnel increased,^9–11^ and vulnerable adults were at high risk of hospitalisation and death. However, BCG vaccines were available. In this context, several randomised placebo-controlled BCG trials with COVID-19 endpoints were initiated during the pandemic to determine whether BCG vaccination reduces SARS-CoV-2 infection, and/or infection severity or duration.

We recently published the results of the BCG-Corona trial in 1,511 Dutch HCWs.^12^ The primary endpoint of this trial was absenteeism for any reason. Secondary endpoints were, among others, participant-reported positive SARS-CoV-2 tests, and symptomatic respiratory infections. None of these endpoints differed statistically significantly between the BCG and placebo groups. However, while participants were instructed to always get tested in case of COVID-19-like symptoms, they may not always have done so, especially in the case of mild infections. Furthermore, asymptomatic infections likely went unnoticed. We therefore also collected blood samples for SARS-CoV-2 antibody testing at 3-6 months and 12 months post-vaccination. This uncovered 74 additional infections in addition to the 224 participant-reported infections. Our aim was to determine whether BCG vaccination compared to placebo vaccination reduces SARS-CoV-2 infection acquisition, severity and/or duration using this most comprehensive dataset in the field to date, with endpoints detected by both self-report and serology and with daily data on symptoms.

## Methods

### Study design and population

The BCG-Corona trial was a multi-centre, double-blind, placebo-controlled randomised trial comparing BCG to placebo vaccination to prevent absenteeism and COVID-19-related endpoints. The study protocol was approved by the institutional review board of the University Medical Center (UMC) Utrecht, registered at clinicaltrials.gov (identifier: NCT04328441), and published.^13^ The sample size was determined for the primary objective (absenteeism) by computer simulation. Participants were doctors, nurses, paramedics, and support staff from 9 Dutch hospitals: three university hospitals that implemented in-hospital sampling (referred to as the core hospitals) and 6 hospitals (one university and 5 non-university teaching hospitals) that did not. Participants were 18 years or older, were expected to be in direct contact with SARS-CoV-2-infected patients, and possessed a smartphone. The primary exclusion criteria were known allergy to BCG, active or latent *Mycobacterium tuberculosis* infection (as judged by the local Principal Investigator in each hospital), any other active infection, immunocompromised state, malignancy, or lymphoma in the past two years, current or planned pregnancy, any vaccination in the past 4 weeks, having a hospital employment contract of less than 22 hours per week, or expected work absence of at least 4 weeks.

### Study procedures

After having obtained informed consent, participants were randomised to BCG or placebo (1:1) using a computer-generated dynamic randomisation algorithm in random blocks of 2, 4, or 6 sequences, stratified by hospital. They received an intradermal injection in the left upper arm with either 0.1 ml of the Danish strain 1331 (Statens Serum Institut, Denmark), equivalent to 0.075 mg attenuated *Mycobacterium bovis*, or 0.1 ml of normal saline solution. Participants and study personnel conducting participant follow-up were blinded to treatment allocation. Study personnel preparing and administering the study vaccines, and data analysts, were not blinded but could not influence treatment allocations or data collection.

At the randomisation visit, participants completed an online baseline questionnaire (Research Online, Julius Center, UMC Utrecht, the Netherlands) and installed a diary application on their smartphone (Research Follow App, Your Research BV, Huizen, Netherlands). Participants were asked to report symptoms via the app daily, and SARS-CoV-2 exposures and test results, and healthcare visits including hospital admissions, weekly. The daily questionnaire was integrated into the weekly questionnaire after 6 months to improve user convenience, and COVID-19 vaccination questions were added after the start of the Dutch vaccination campaign on 6 January 2021. We used push notifications, emails, and phone calls to maximise adherence with app completion, and we terminated the app for all participants on 27 March 2021. Blood sampling continued until mid-June 2021, and we collected data on symptoms, positive SARS-CoV-2 tests, and COVID-19 vaccinations for the period between 27 March 2021 and the participant’s final sampling date via an online questionnaire (Formdesk, Innovero Software Solutions BV, Wassenaar, Netherlands) and email.

Blood samples were collected in two sampling rounds, dividing the follow-up time into two potential seroconversion periods. Period 1 is the period between study vaccination and the first sampling round, after about three months (M3) for the core hospitals and about 6 months (M6) for the other hospitals. Period 2 is the period between the first and the second sampling rounds, about 12 months (M12) after study vaccination for all hospitals. Core hospital participants donated 10 ml serum via in-hospital venepuncture, which was processed, frozen, and transported frozen to the Dutch National Institute for Public Health and the Environment (RIVM in Dutch). The other participants, as well as core hospital participants who missed their venepuncture visit, were asked to collect about 300 uL peripheral capillary blood at home by fingerprick using a sampling kit that was sent to them by mail; they returned the sample by mail to the RIVM laboratory on the day of collection. The RIVM laboratory used an in-house magnetic immunoassay on a Luminex platform to determine the presence and concentrations of SARS-CoV-2-specific antibodies (supplementary methods).^14–16^ At the end of period 1, only immunoglobulin G (IgG) antibodies against the S1 subunit of the SARS-CoV-2 spike protein (anti-S1) were measured. At the end of period 2, both anti-S1 and IgG antibodies against the SARS-CoV-2 nucleocapsid protein (anti-N) were measured to enable differentiation between natural infections and COVID-19 vaccine-induced antibodies.

### Outcome and follow-up time definitions

The presence of a SARS-CoV-2 infection was defined as a participant-reported positive test result and/or seroconversion, which in turn was defined as anti-S1 seropositivity at the end of period 1 or anti-S1 and anti-N seropositivity at the end of period 2 (supplementary methods). Participants with anti-N but no anti-S1 seropositivity at the end of period 2, and no corresponding positive test-date, were classified as having ‘inconclusive episodes’ and were excluded from most analyses except some sensitivity analyses (supplement). Participants who had less than 80% app completion without self-reported or serological evidence of an infection were also excluded because we cannot be sure that they are true negatives.

Episode duration and severity were based on the diary app data (supplementary methods). The acute infection episode duration was defined as the number of consecutive days during which the participant reported symptoms, not including standalone loss of smell/taste or lingering symptoms (e.g. fatigue) after respiratory symptoms had ceased. Fever was defined as a temperature of 38°C or above. All other symptoms were reported on a scale of 0-5: 0 for not present and 1-5 corresponding to increasing severity. Episode severity was categorised as asymptomatic, mild, or mild-to-moderate using both episode duration and symptoms severity data (definitions in Table S1). The three hospitalisations due to COVID-19 were included in the mild-to-moderate category. Separate variables were created for long-term loss of smell/taste and long-COVID (based on symptoms other than standalone loss of smell/taste). These were defined as continuing to report the respective symptoms for at least 60 days after the end of the acute infection episode.

The number of follow-up days was calculated as the number of days between vaccination and the first infection episode (the positive test-date or the start-date of the symptomatic period if no test-date available; survival analyses) or the end of study (the M12 sampling date or the last day of app completion if the participant did not provide a M12 sample; all other analyses). The app completion percentage was calculated for the period between vaccination and 27 March 2021 (supplementary methods).

### Statistical analyses

Analyses were performed in R version 4.1.2 (PBC, Boston, MA, USA). We compared characteristics between randomisation groups using Wilcoxon rank sum test for continuous variables and Chi-squared test for categorical variables. Endpoints were any SARS-CoV-2 infection, or asymptomatic, mild or mild-to-moderate infections; only the first infection per participant was included. The occurrence of endpoints in the randomisation groups was modelled using (multinomial) logistic regression models for cumulative incidence and Cox proportional hazards models for time to first event (the latter including infections that could be dated only). All models were first run unadjusted, followed by adjustment for potential confounders. Randomisation group was forced into the multivariable models, and potential confounders were selected using a backward stepwise approach using the Akaike Information Criterion (k=2.7) to select the final model. Sensitivity analyses were conducted assuming that the inconclusive episodes were either true episodes or not, or assuming that participants with less than 80% app completion and no reported or detected infection episode were true negatives.

## Results

### Participant flow and baseline characteristics

Between 24 March and 23 April 2020, 1,526 HCWs were screened and 1,511 randomised: 753 to the BCG and 758 to the placebo group (Figure 1). Participants with less than 80% app completion and no evidence of infection (Figure S1), or with inconclusive episodes only, were removed (68 and 20 in the BCG group and 98 and 16 in the placebo group, respectively). The analysis population therefore consisted of 1,309 participants: 665 in the BCG group and 644 in the placebo group. Of the analysis population, 82.9% in the BCG group and 85.7% in the placebo group provided a blood sample in the second sampling round. An additional 9.6% and 9.5%, respectively, provided a blood sample in the first but not in the second sampling round. None of the data availability characteristics differed significantly between the randomisation groups (Figure 1).

**Figure 1:**
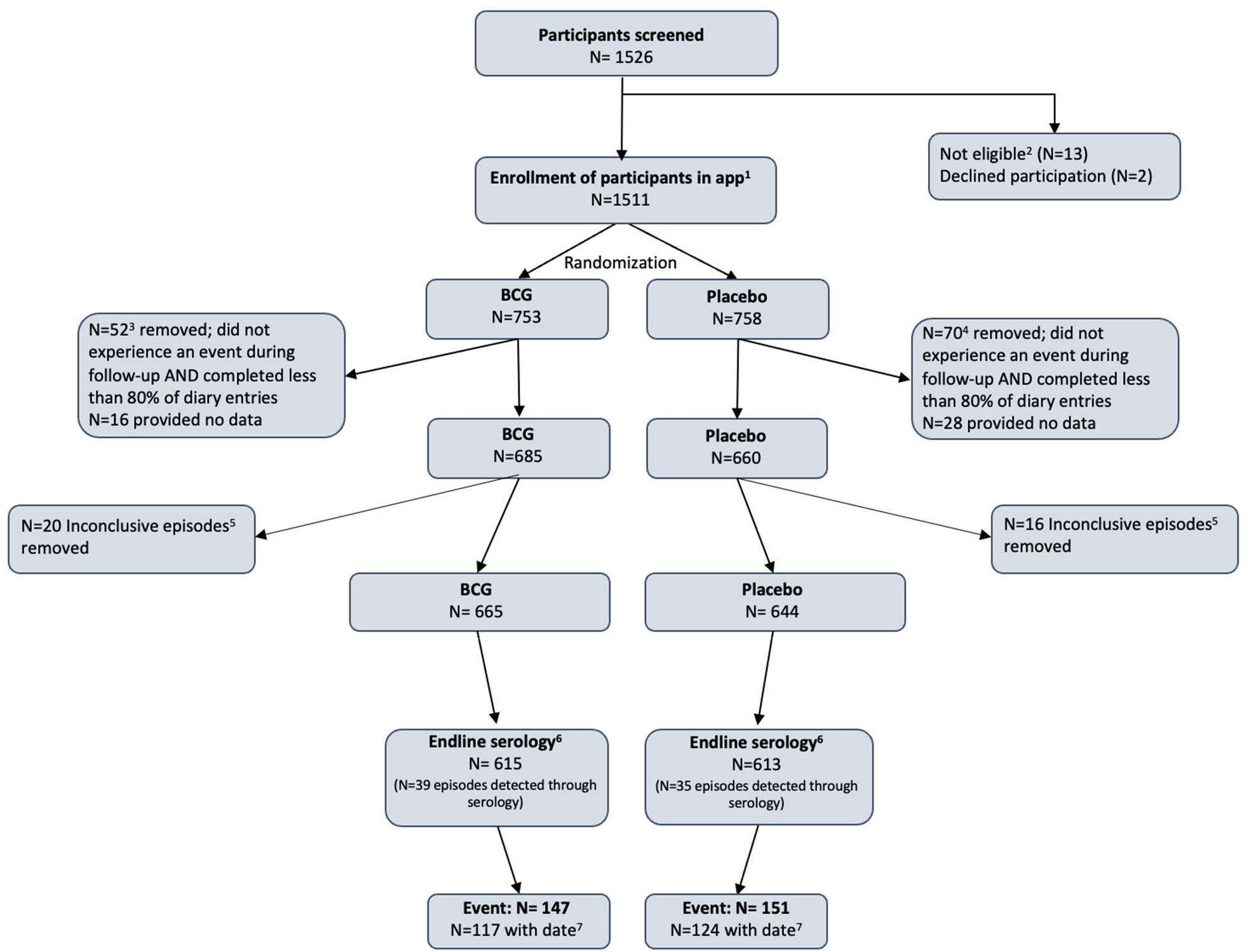
BCG-Corona Study Flow. 1. The first participant was enrolled on 24 March 2020 and the last participant on 23 April 2020. The last day of follow up in the app was 27 March 2021, with additional participant information obtained via an online questionnaire to cover the period between 28 March 2021 and the last blood sampling date, if applicable. 2. Thirteen participants were not eligible: pregnancy (N=4), severely immunocompromised state (N=1), malignancy/ lymphoma in the last two years (N=1), active viral/bacterial infection (N=2), recent or expected vaccination (N=3), not hospital personnel caring for COVID-19 patient (N=1), or *Mycobacterium tuberculosis* infection history (N=1). 3. Of the 52 participants in the BCG group removed due to less than 80% diary app completion and no evidence of infection, 19 terminated study participation early at their own request, 10 became complete non-responders prior to the end of the study for unknown reasons, two went on pregnancy leave, and 21 entered data inconsistently. An additional 16 participants never entered any data in the app. 4. Of the 70 participants in the placebo group removed due to less than 80% diary app completion and no evidence of infection, 33 terminated study participation early at their own request, 10 became complete non-responders prior to the end of the study for unknown reasons, 6 terminated hospital employment, three went on pregnancy leave, and 18 entered data inconsistently. An additional 28 participants never entered any data in the app. The percentage of participants that completed less than 80% of the diary entries in the BCG group was not statistically different from the percentage in the placebo group (Chi-squared p=0.283). 5. An inconclusive episode is defined as anti-N seropositivity but not anti-S1 seropositivity at the end of period 2. The percentage of participants with inconclusive episodes only removed in the BCG group, was not statistically different from the percentage in the placebo group (Chi-squared p=0.599). 6. N is the number of participants that participated in at least one sampling round at the end of the follow-up (Chi-squared p=0.237). 7. Events with a date were included in all analyses and events without a date in logistic regressions only.

The majority of the 1,309 participants (74.4%) were female, with no difference between groups (Table 1). The age of participants ranged between 18-67 years, with mean ages of 41.8 years in the BCG group and 43.2 years in the placebo group. Job-related characteristics were also similar between groups: 49.3% of the participants were nurses and 53.1% reported to have direct patient contact at least 75% of their work hours. Household size, smoking behaviour, histories of BCG or recent influenza or other vaccinations, ever having had a positive tuberculosis test, having had a respiratory tract infection last winter, and chronic comorbidities were well-balanced between groups. The baseline characteristics of the analysis population were comparable to those of the entire randomised population (Table S2). About half of the participants (47.2% in the BCG group and 50.6% in the placebo group; p=0.218) received at least one dose of one of 5 different COVID-19 vaccines between 6 January 2021 and the end of follow-up. The time to first COVID-19 vaccination did not differ between the randomisation groups (Figure 2).

**Table 1:**
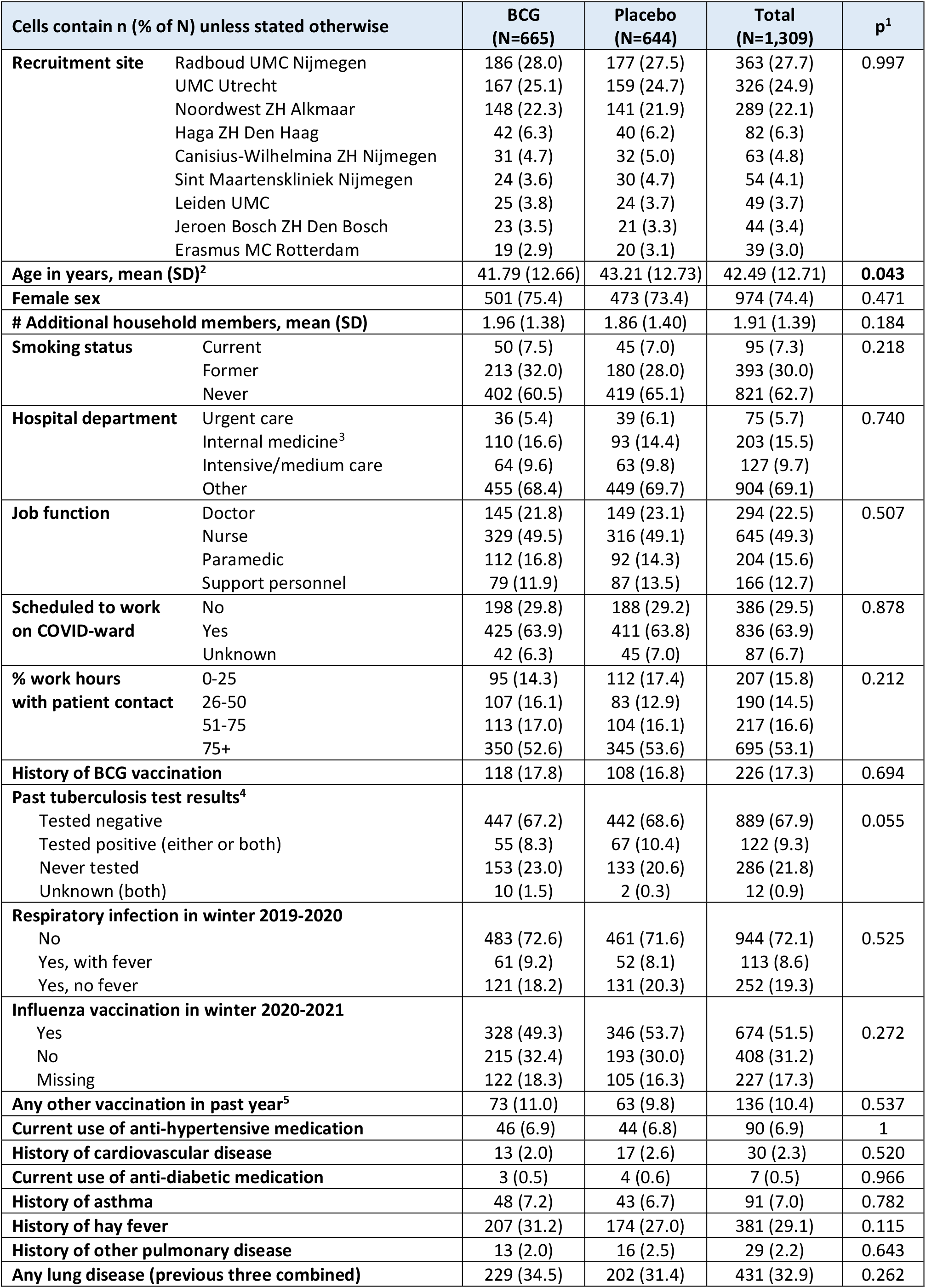
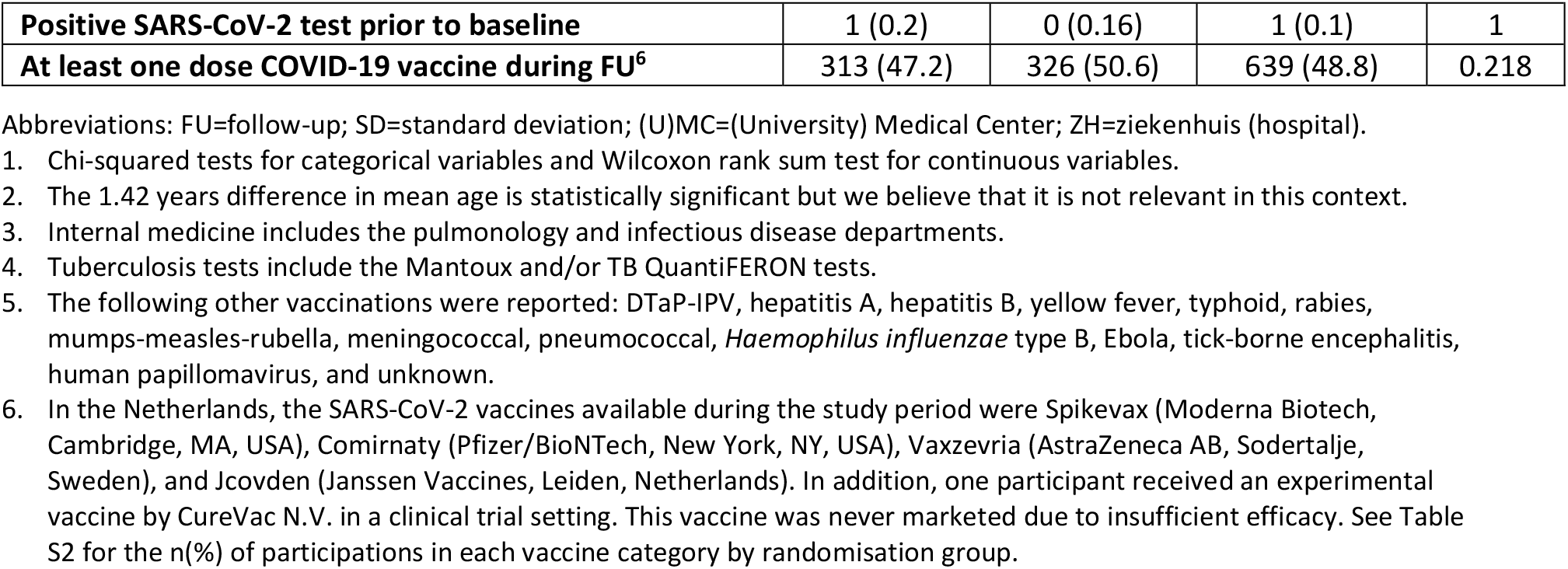
Baseline characteristics of the analysis population by study group.

| Cells contain n (% of N) unless stated otherwise |  | BCG<br>(N=665) | Placebo<br>(N=644) | Total<br>(N=1,309) | p<sup>1</sup> |
| --- | --- | --- | --- | --- | --- |
| **Recruitment site** | Radboud UMC Nijmegen | 186 (28.0) | 177 (27.5) | 363 (27.7) | 0.997 |
|  | UMC Utrecht | 167 (25.1) | 159 (24.7) | 326 (24.9) |  |
|  | Noordwest ZH Alkmaar | 148 (22.3) | 141 (21.9) | 289 (22.1) |  |
|  | Haga ZH Den Haag | 42 (6.3) | 40 (6.2) | 82 (6.3) |  |
|  | Canisius-Wilhelmina ZH Nijmegen | 31 (4.7) | 32 (5.0) | 63 (4.8) |  |
|  | Sint Maartenskliniek Nijmegen | 24 (3.6) | 30 (4.7) | 54 (4.1) |  |
|  | Leiden UMC | 25 (3.8) | 24 (3.7) | 49 (3.7) |  |
|  | Jeroen Bosch ZH Den Bosch | 23 (3.5) | 21 (3.3) | 44 (3.4) |  |
|  | Erasmus MC Rotterdam | 19 (2.9) | 20 (3.1) | 39 (3.0) |  |
| **Age in years, mean (SD)<sup>2</sup>** |  | 41.79 (12.66) | 43.21 (12.73) | 42.49 (12.71) | **0.043** |
| **Female sex** |  | 501 (75.4) | 473 (73.4) | 974 (74.4) | 0.471 |
| **# Additional household members, mean (SD)** |  | 1.96 (1.38) | 1.86 (1.40) | 1.91 (1.39) | 0.184 |
| **Smoking status** | Current | 50 (7.5) | 45 (7.0) | 95 (7.3) | 0.218 |
|  | Former | 213 (32.0) | 180 (28.0) | 393 (30.0) |  |
|  | Never | 402 (60.5) | 419 (65.1) | 821 (62.7) |  |
| **Hospital department** | Urgent care | 36 (5.4) | 39 (6.1) | 75 (5.7) | 0.740 |
|  | Internal medicine<sup>3</sup> | 110 (16.6) | 93 (14.4) | 203 (15.5) |  |
|  | Intensive/medium care | 64 (9.6) | 63 (9.8) | 127 (9.7) |  |
|  | Other | 455 (68.4) | 449 (69.7) | 904 (69.1) |  |
| **Job function** | Doctor | 145 (21.8) | 149 (23.1) | 294 (22.5) | 0.507 |
|  | Nurse | 329 (49.5) | 316 (49.1) | 645 (49.3) |  |
|  | Paramedic | 112 (16.8) | 92 (14.3) | 204 (15.6) |  |
|  | Support personnel | 79 (11.9) | 87 (13.5) | 166 (12.7) |  |
| **Scheduled to work on COVID-ward** | No | 198 (29.8) | 188 (29.2) | 386 (29.5) | 0.878 |
|  | Yes | 425 (63.9) | 411 (63.8) | 836 (63.9) |  |
|  | Unknown | 42 (6.3) | 45 (7.0) | 87 (6.7) |  |
| **% work hours with patient contact** | 0-25 | 95 (14.3) | 112 (17.4) | 207 (15.8) | 0.212 |
|  | 26-50 | 107 (16.1) | 83 (12.9) | 190 (14.5) |  |
|  | 51-75 | 113 (17.0) | 104 (16.1) | 217 (16.6) |  |
|  | 75+ | 350 (52.6) | 345 (53.6) | 695 (53.1) |  |
| **History of BCG vaccination** |  | 118 (17.8) | 108 (16.8) | 226 (17.3) | 0.694 |
| **Past tuberculosis test results<sup>4</sup>** |  |  |  |  | 0.055 |
| Tested negative |  | 447 (67.2) | 442 (68.6) | 889 (67.9) |  |
| Tested positive (either or both) |  | 55 (8.3) | 67 (10.4) | 122 (9.3) |  |
| Never tested |  | 153 (23.0) | 133 (20.6) | 286 (21.8) |  |
| Unknown (both) |  | 10 (1.5) | 2 (0.3) | 12 (0.9) |  |
| **Respiratory infection in winter 2019-2020** | No | 483 (72.6) | 461 (71.6) | 944 (72.1) | 0.525 |
|  | Yes, with fever | 61 (9.2) | 52 (8.1) | 113 (8.6) |  |
|  | Yes, no fever | 121 (18.2) | 131 (20.3) | 252 (19.3) |  |
| **Influenza vaccination in winter 2020-2021** | Yes | 328 (49.3) | 346 (53.7) | 674 (51.5) | 0.272 |
|  | No | 215 (32.4) | 193 (30.0) | 408 (31.2) |  |
|  | Missing | 122 (18.3) | 105 (16.3) | 227 (17.3) |  |
| **Any other vaccination in past year<sup>5</sup>** |  | 73 (11.0) | 63 (9.8) | 136 (10.4) | 0.537 |
| **Current use of anti-hypertensive medication** |  | 46 (6.9) | 44 (6.8) | 90 (6.9) | 1 |
| **History of cardiovascular disease** |  | 13 (2.0) | 17 (2.6) | 30 (2.3) | 0.520 |
| **Current use of anti-diabetic medication** |  | 3 (0.5) | 4 (0.6) | 7 (0.5) | 0.966 |
| **History of asthma** |  | 48 (7.2) | 43 (6.7) | 91 (7.0) | 0.782 |
| **History of hay fever** |  | 207 (31.2) | 174 (27.0) | 381 (29.1) | 0.115 |
| **History of other pulmonary disease** |  | 13 (2.0) | 16 (2.5) | 29 (2.2) | 0.643 |
| **Any lung disease (previous three combined)** |  | 229 (34.5) | 202 (31.4) | 431 (32.9) | 0.262 |

|  |  |  |  |  |
| --- | --- | --- | --- | --- |
| **Positive SARS-CoV-2 test prior to baseline** | **1 (0.2)** | **0 (0.16)** | **1 (0.1)** | **1** |
| **At least one dose COVID-19 vaccine during FU<sup>6</sup>** | **313 (47.2)** | **326 (50.6)** | **639 (48.8)** | **0.218** |

Abbreviations: FU=follow-up; SD=standard deviation; (U)MC=(University) Medical Center; ZH=ziekenhuis (hospital).

**Figure 2:**
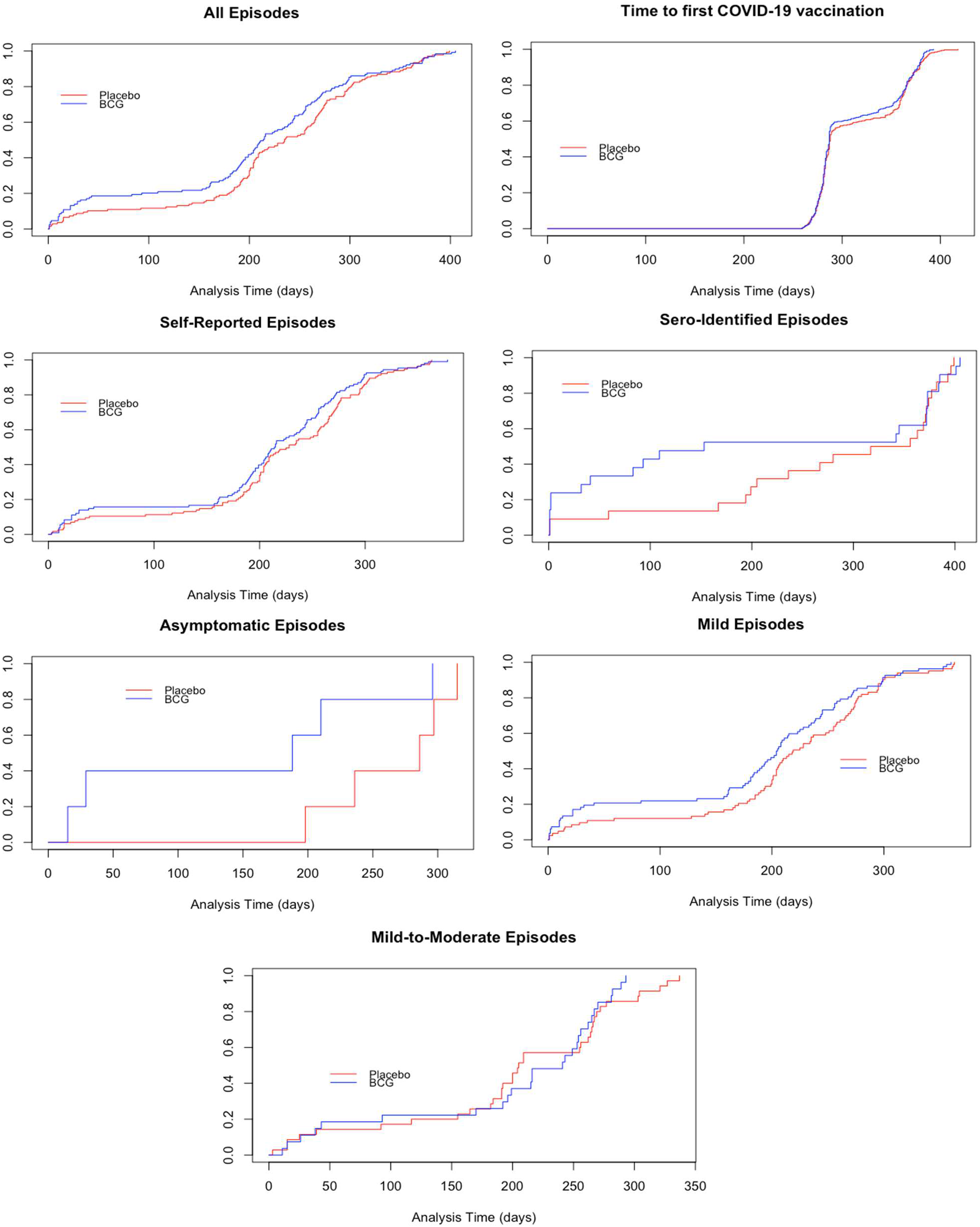
Time to first event curves. 1. Participant-reported episodes include proven episodes with and without seroconversion. 2. Sero-identified episodes includes all possible episodes detected through serology that could be linked to a symptomatic period and therefore have a known episode infection-date.

### Descriptive outcomes by randomisation group

SARS-CoV-2 infections over calendar time in the study population mirrored epidemic waves in the Netherlands (Figure S2). Only one participant (in the BCG group) reported to have had a positive SARS-CoV-2 test prior to randomisation. A total of 298 infections (not including 4 second infections) were detected during the trial, of which 224 were participant-reported and 74 detected by serology only (Table 2). Overall, 147/665 participants (22.1%) in the BCG group and 151/644 (23.4%) participants in the placebo group experienced an infection during follow-up (p=0.608). The SARS-CoV-2 incidence was 0.25 per person-year in the BCG group and 0.26 per person-year in the placebo group (incidence rate ratio=0.95; 95% CI 0.76-1.21; p=0.732) (Table S3).

**Table 2:**
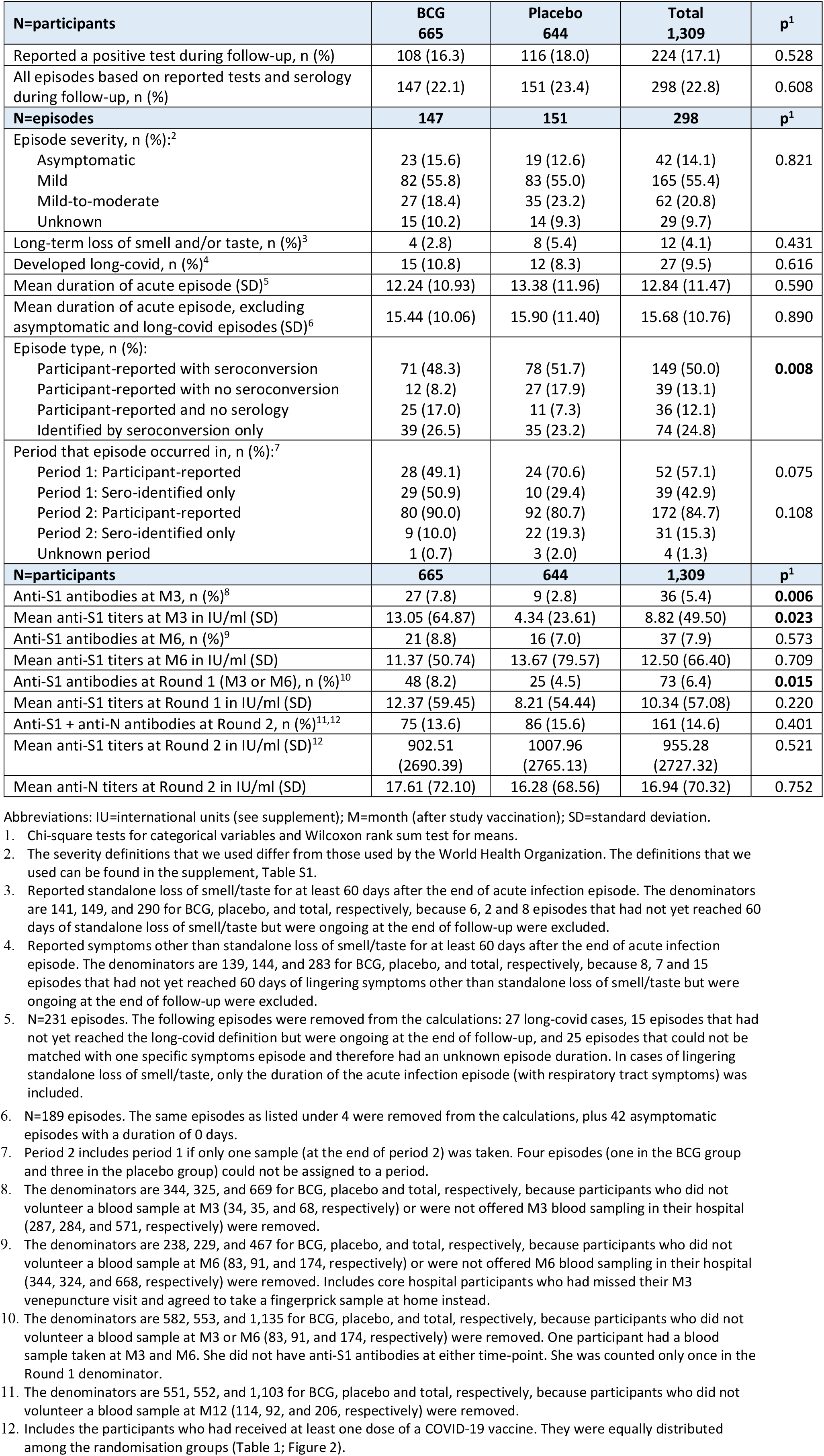
Outcomes during follow-up by randomisation group.

In the BCG and placebo groups, respectively, 18.4% and 23.2% were mild-to-moderate, 55.8% and 55.0% were mild, and 15.6% and 12.6% were asymptomatic (no statistically significant differences; Table 2). As expected, most symptomatic infections were identified via participant-reporting: 90.3% of mild and 96.8% of mild-to-moderate episodes. In contrast, 76.2% of the asymptomatic episodes were identified by serology only. Fifteen episodes in the BCG group (10.8%) and 12 in the placebo group (8.3%) reached the long-COVID definition during follow-up (p=0.616). In addition, 4 (2.8%) and 8 (5.4%) episodes in the BCG and placebo groups, respectively, resulted in long-term loss of smell/taste (p=0.431) (Table 2). The mean episode durations of acute episodes (not including long-COVID or asymptomatic episodes) were 15.4 days (standard deviation (SD)=10.1 days) in the BCG group and 15.9 days (SD=11.4 days) in the placebo group (p=0.890; Table 2, Figure S3). None of the different types of symptoms, nor their individual severities, differed between randomisation groups (Table S3).

About a quarter of all infections (24.8%) was identified by serology only, and this percentage was non-significantly higher in the BCG group in period 1 (50.9%) compared to the BCG group in period 2 (10.0%) and the placebo group in both periods (29.4% and 19.3%; Table 2). The percentage of participants with seroconversion was higher in the BCG than in the placebo group at M3 (7.8% versus 2.8%; p=0.006), but not at M6 (8.8% versus 7.0%; p=0.573) or M12 (13.6% versus 15.6%; p=0.401).

Similarly, the mean anti-S1 antibody concentration was higher in the BCG than in the placebo group at M3, but there were no differences in anti-S1 or anti-N antibody concentrations between the groups at M6 or M12 (Table 2).

### Logistic regression and Cox proportional hazards models

Unadjusted and adjusted logistic regression models showed no differences between BCG and placebo vaccination in cumulative incidence of any SARS-CoV-2 infection or of asymptomatic, mild, or mild-to-moderate infections separately (Table 3). No differences were seen in Cox proportional hazards time-to-first-event models with these same outcomes either (Table 3, Figure 2). Younger age, various job-related characteristics, past BCG vaccination, and current use of hypertension medication were associated with increased odds or hazards of acquiring infection (Table S4) and were retained in all multivariable models (Table 3, Table S5). A larger household size was associated with increased odds or hazards in some but not all models. In terms of job-related characteristics, being a nurse or support staff (compared to a doctor), having a larger percentage of work hours with patient contact, and expectation to work on a COVID-ward were associated with higher odds or hazards of infection or infection severity, and working in the intensive or medium care departments (compared to the urgent care department) with lower odds or hazards. Recruitment site, enrolment week, sex, ever having tested positive for tuberculosis, current use of antidiabetic medication, history of pulmonary disease, and history of cardiovascular disease were not associated with an outcome in any of the models. All sensitivity analyses showed similar results (Tables S6-S7).

**Table 3:**
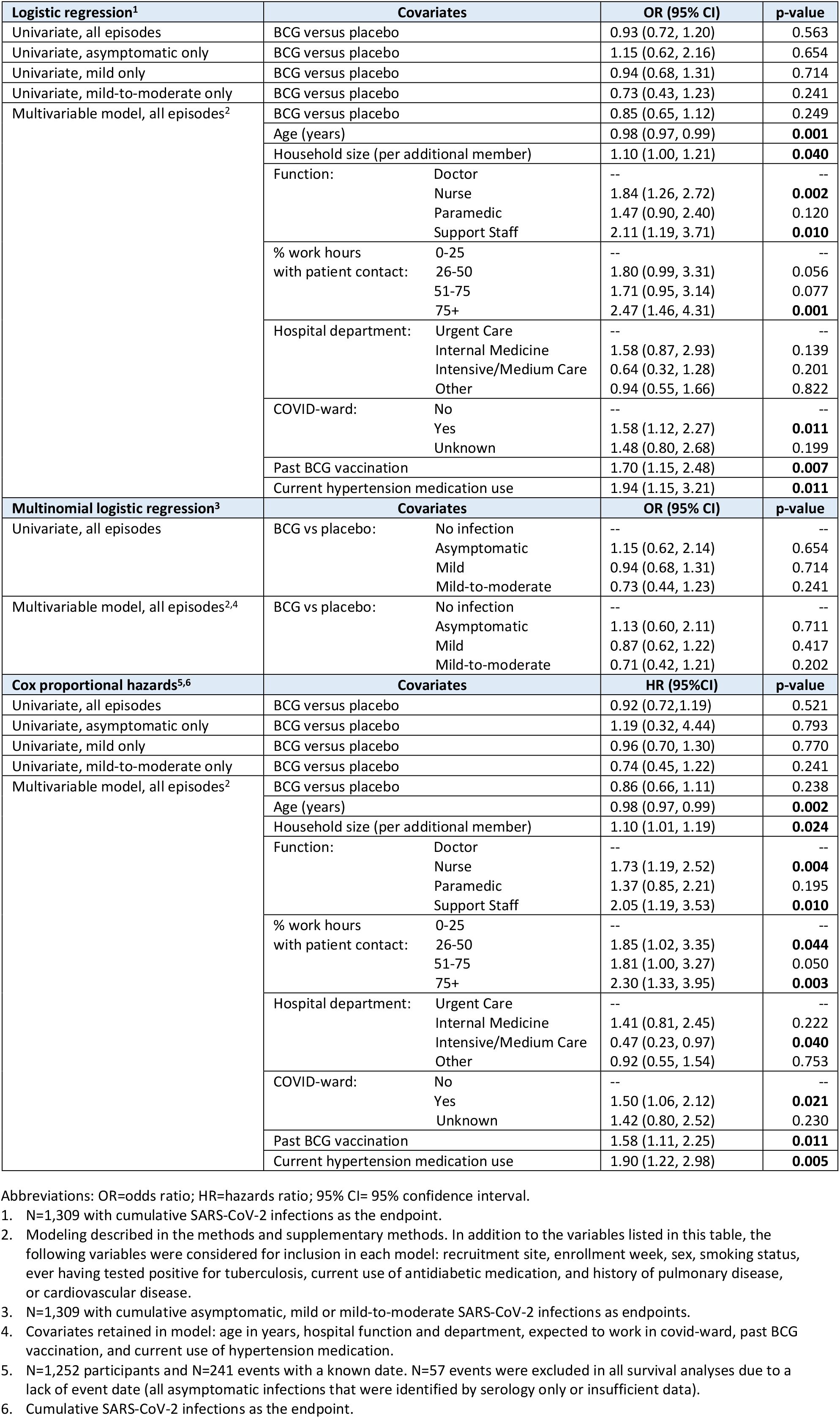
Logistic regression and Cox proportional hazards models.

| **Logistic regression<sup>1</sup>** | **Covariates** | **OR (95% CI)** | **p-value** |
| --- | --- | --- | --- |
| Univariate, all episodes | BCG versus placebo | 0.93 (0.72, 1.20) | 0.563 |
| Univariate, asymptomatic only | BCG versus placebo | 1.15 (0.62, 2.16) | 0.654 |
| Univariate, mild only | BCG versus placebo | 0.94 (0.68, 1.31) | 0.714 |
| Univariate, mild-to-moderate only | BCG versus placebo | 0.73 (0.43, 1.23) | 0.241 |
| Multivariable model, all episodes<sup>2</sup> | BCG versus placebo | 0.85 (0.65, 1.12) | 0.249 |
|  | Age (years) | 0.98 (0.97, 0.99) | **0.001** |
|  | Household size (per additional member) | 1.10 (1.00, 1.21) | **0.040** |
|  | Function: Doctor | -- | -- |
|  | Nurse | 1.84 (1.26, 2.72) | **0.002** |
|  | Paramedic | 1.47 (0.90, 2.40) | 0.120 |
|  | Support Staff | 2.11 (1.19, 3.71) | **0.010** |
|  | % work hours with patient contact: 0-25 | -- | -- |
|  | 26-50 | 1.80 (0.99, 3.31) | 0.056 |
|  | 51-75 | 1.71 (0.95, 3.14) | 0.077 |
|  | 75+ | 2.47 (1.46, 4.31) | **0.001** |
|  | Hospital department: Urgent Care | -- | -- |
|  | Internal Medicine | 1.58 (0.87, 2.93) | 0.139 |
|  | Intensive/Medium Care | 0.64 (0.32, 1.28) | 0.201 |
|  | Other | 0.94 (0.55, 1.66) | 0.822 |
|  | COVID-ward: No | -- | -- |
|  | Yes | 1.58 (1.12, 2.27) | **0.011** |
|  | Unknown | 1.48 (0.80, 2.68) | 0.199 |
|  | Past BCG vaccination | 1.70 (1.15, 2.48) | **0.007** |
|  | Current hypertension medication use | 1.94 (1.15, 3.21) | **0.011** |
| **Multinomial logistic regression<sup>3</sup>** | **Covariates** | **OR (95% CI)** | **p-value** |
| Univariate, all episodes | BCG vs placebo: No infection | -- | -- |
|  | Asymptomatic | 1.15 (0.62, 2.14) | 0.654 |
|  | Mild | 0.94 (0.68, 1.31) | 0.714 |
|  | Mild-to-moderate | 0.73 (0.44, 1.23) | 0.241 |
| Multivariable model, all episodes<sup>2,4</sup> | BCG vs placebo: No infection | -- | -- |
|  | Asymptomatic | 1.13 (0.60, 2.11) | 0.711 |
|  | Mild | 0.87 (0.62, 1.22) | 0.417 |
|  | Mild-to-moderate | 0.71 (0.42, 1.21) | 0.202 |
| **Cox proportional hazards<sup>5,6</sup>** | **Covariates** | **HR (95%CI)** | **p-value** |
| Univariate, all episodes | BCG versus placebo | 0.92 (0.72,1.19) | 0.521 |
| Univariate, asymptomatic only | BCG versus placebo | 1.19 (0.32, 4.44) | 0.793 |
| Univariate, mild only | BCG versus placebo | 0.96 (0.70, 1.30) | 0.770 |
| Univariate, mild-to-moderate only | BCG versus placebo | 0.74 (0.45, 1.22) | 0.241 |
| Multivariable model, all episodes<sup>2</sup> | BCG versus placebo | 0.86 (0.66, 1.11) | 0.238 |
|  | Age (years) | 0.98 (0.97, 0.99) | **0.002** |
|  | Household size (per additional member) | 1.10 (1.01, 1.19) | **0.024** |
|  | Function: Doctor | -- | -- |
|  | Nurse | 1.73 (1.19, 2.52) | **0.004** |
|  | Paramedic | 1.37 (0.85, 2.21) | 0.195 |
|  | Support Staff | 2.05 (1.19, 3.53) | **0.010** |
|  | % work hours with patient contact: 0-25 | -- | -- |
|  | 26-50 | 1.85 (1.02, 3.35) | **0.044** |
|  | 51-75 | 1.81 (1.00, 3.27) | 0.050 |
|  | 75+ | 2.30 (1.33, 3.95) | **0.003** |
|  | Hospital department: Urgent Care | -- | -- |
|  | Internal Medicine | 1.41 (0.81, 2.45) | 0.222 |
|  | Intensive/Medium Care | 0.47 (0.23, 0.97) | **0.040** |
|  | Other | 0.92 (0.55, 1.54) | 0.753 |
|  | COVID-ward: No | -- | -- |
|  | Yes | 1.50 (1.06, 2.12) | **0.021** |
|  | Unknown | 1.42 (0.80, 2.52) | 0.230 |
|  | Past BCG vaccination | 1.58 (1.11, 2.25) | **0.011** |
|  | Current hypertension medication use | 1.90 (1.22, 2.98) | **0.005** |

Abbreviations: OR=odds ratio; HR=hazards ratio; 95% CI= 95% confidence interval.
1. N=1,309 with cumulative SARS-CoV-2 infections as the endpoint. 2. Modeling described in the methods and supplementary methods. In addition to the variables listed in this table, the following variables were considered for inclusion in each model: recruitment site, enrollment week, sex, smoking status, ever having tested positive for tuberculosis, current use of antidiabetic medication, and history of pulmonary disease, or cardiovascular disease. 3. N=1,309 with cumulative asymptomatic, mild or mild-to-moderate SARS-CoV-2 infections as endpoints. 4. Covariates retained in model: age in years, hospital function and department, expected to work in covid-ward, past BCG vaccination, and current use of hypertension medication. 5. N=1,252 participants and N=241 events with a known date. N=57 events were excluded in all survival analyses due to a lack of event date (all asymptomatic infections that were identified by serology only or insufficient data). 6. Cumulative SARS-CoV-2 infections as the endpoint.

## Discussion

This is the most comprehensive analysis of a placebo-controlled BCG trial with SARS-CoV-2 endpoints to date, including both participant-reported positive tests as well as serology to identify infections and in-depth characterisation of infection episodes using daily symptoms data. Serology detected an additional 74 endpoints on top of the 224 participant-reported endpoints. Unadjusted and adjusted logistic regression (cumulative incidence) and Cox proportional hazards (time-to-first-event) models showed no differences between BCG and placebo vaccination for SARS-CoV-2 infections of any severity, nor for asymptomatic, mild, or mild-to-moderate infections separately. Mean infection durations, and the proportions of participants with infections who developed long-COVID or long-term loss of smell/taste, did not differ between the randomisation groups either.

Our findings corroborate the results of several placebo-controlled BCG trials with SARS-CoV-2 endpoints. A South African trial in HCWs (N=1,000),^17^ and two Dutch trials in community-dwelling (N=2,014)^18^ and vulnerable elderly populations (N=6,112),^19^ found no differences between BCG and placebo vaccination in participant-reported positive SARS-CoV-2 tests, COVID-19 hospitalisations or deaths, or any symptomatic respiratory tract infection, in the year following vaccination. The South African trial did not detect a difference in the cumulative incidence of SARS-CoV-2 seroconversion either, but it is unclear how seroconversion was defined, and seroconversions due to natural infection versus COVID-19 vaccination were not differentiated.^17^ The ACTIVATE-2 trial in older Greek patients (N=153) did report a protective effect of BCG vaccination (odds ratio (OR) 0.32, 95% confidence interval (CI) 0.13-0.79) but the trial endpoint combined test-confirmed and suspected COVID-19 cases.^20^ A trial in India (N=495) reported a protective effect for suspected COVID-19 cases (OR 0.38, 95% CI 0.20–0.72) but not for test-confirmed cases (OR 1.08, 95% CI 0.54–2.14).^21^ It has been hypothesised that childhood BCG vaccination and/or latent tuberculosis might modulate BCG-induced trained immunity.^22^ Childhood vaccination is common in South Africa, Greece, and India but not in the Netherlands. Latent tuberculosis is more prevalent in South Africa than in Greece (48.5% and 5.8% in the South African and Greek trial populations, respectively),^4,17^ and uncommon in the Netherlands. It therefore seems unlikely that these factors explain the divergent results of the Greek and Indian versus South African and Dutch trials, but ongoing trials in additional populations might shed more light on this debate in the future.^23,24^ We think that it is more likely that endpoints based on suspected COVID-19 cases are insufficiently specific for SARS-CoV-2 infection, and that the total number of endpoints in the Greek and Indian trials were too small to rule out chance.^20,21^

SARS-CoV-2 risk in HCWs increased with the proportion of work hours with patient contact. This suggests that the risk is mostly due to a high probability of exposure rather than a higher risk of infection once exposed. Interestingly, working in intensive or medium care was associated with a lower risk of infection than working in urgent care, which may be related to better infection control measures, including availability of personal protective equipment, in the former. Using antihypertensive medication was also consistently associated with higher infection risk. Patients diagnosed with high blood pressure have increased levels of angiotensin-converting enzyme 2 (ACE2), which acts as the entry receptor for SARS-CoV-2.^25,26^ Whether antihypertensive medication use has an additional effect is controversial: animal models have suggested an increase in ACE2 in the respiratory tract due to these medications but an increased SARS-CoV-2 infection risk after medication initiation has not been confirmed in humans.^27,28^ Finally, our data confirm that a substantial proportion of SARS-CoV-2 infections (14.1%) are asymptomatic, and that HCWs are less likely to seek testing for asymptomatic or mild infections than for mild-to-moderate infections.

The BCG-Corona trial was initiated in March 2020, early in the first epidemic wave in the Netherlands, and therefore captured SARS-CoV-2 exposures and infections in an immunologically naïve population. This may explain why the mean acute infection episode duration was long (15.7 days) and a substantial proportion of infected individuals developed lingering symptoms (10% long-COVID plus an additional 4% long-term loss of smell/taste). Studies in the United Kingdom showed that median episode duration declined as the pandemic progressed, from a median of 11 days in 2020,^29^ to 8 days during Delta-dominance in 2021 and 5 days during Omicron-dominance in the Winter of 2021/2022.^30^ The proportion of COVID-19 patients developing long-COVID varies widely between studies due to differences in definitions used. A large population-based cohort in Groningen province in the Netherlands reported a prevalence of 12.7% for the period March 2020 to August 2021 using 23 symptoms including loss of smell/taste and a total symptoms duration of at least 90 days after the diagnosis.^31^

The percentage of participants with SARS-CoV-2 seroconversion at M3 (7.8% versus 2.8%; p=0.006), as well as mean SARS-CoV-2 anti-S1 antibody concentrations (13.1 versus 4.3 IU/ml; p=0.023), were higher in the BCG than in the placebo group, but no differences were seen at M6 or M12. Mean SARS-CoV-2 anti-N antibody concentrations at M12 (not assessed at M3 and M6) also did not differ between randomisation groups. This suggests that BCG vaccination may enhance SARS-CoV-2 antibody production after SARS-CoV-2 infection, but that the effect is short-lived. This finding is not robust because the number of participants with a SARS-CoV-2 infection in the first 3 months of the study was small and the overall anti-S1 antibody concentrations at M3 low. However, additional evidence for BCG potentially acting as an adjuvant comes from human challenge studies in which participants received BCG (re)vaccination followed by influenza or COVID-19 vaccination.^7,32^

Important strengths of this trial are the large number and comprehensive nature of the endpoints. Anti-S1 antibodies are highly sensitive and specific for the presence of the SARS-CoV-2 Spike protein but are induced by both natural infection as well as COVID-19 vaccines.^33,34^ Anti-N seropositivity is considered less sensitive and specific,^33,35^ but is currently the most reliable way to identify natural SARS-CoV-2 infections after COVID-19 vaccination. A recent RIVM study using the same assay that we used in this trial showed that anti-N seropositivity was 85% sensitive for mild infections and 67% sensitive for asymptomatic infections.^16^ While we may have missed some natural infections in the second period of the study, there were no differences between the randomisation groups in the rate of COVID-19 vaccinations, and proportion of participants vaccinated by the end of the study. We would therefore expect a similar number of natural infections to have been missed in each group.

Additional strengths of the trial included the high diary app completion and retention rates. Furthermore, the reliability of participant-reported positive tests was high. The concordance of participant-reported positive test results with hospital laboratory data was 89% in the one hospital for which laboratory data were available (supplementary methods). Negative test results, on the other hand, were substantially underreported. While these were not endpoints in any of our analyses, we did use them to rule out infection episodes potentially responsible for seroconversion. Another potential limitation is that we used two different methods of blood collection: clinician-performed venepuncture and participant-performed fingerprick sampling.^36^

In conclusion, BCG vaccination of HCWs did not reduce SARS-CoV-2 infections nor infection duration or severity (on a scale from asymptomatic to moderate). In the first 3 months after vaccination, BCG vaccination may enhance SARS-CoV-2 antibody production during SARS-CoV-2 infection, but this remains to be confirmed.

### BCG-Corona study group

In addition to the writing team, the BCG-Corona study group also includes (in alphabetical order): Wim G. Boersma (NWZ Alkmaar), Özlem Bulut (Radboud), Reinout van Crevel (Radboud), Priya A. Debisarun (Radboud), Jacobien Hoogerwerf (Radboud), Marien de Jonge (Radboud), Angele P.M. Kerckhoffs (JBZ Den Bosch), Edward Knol (UMC Utrecht), Jan Pieter R. Koopman (Leiden UMC), Vincent P. Kuiper (Leiden UMC), Arief Lalmohamed (UMC Utrecht), Simone J.C.F.M. Moorlag (Radboud), Stephan Nierkens (UMC Utrecht), Cees van Nieuwkoop (HZ Den Haag), Jaap ten Oever (Radboud), Tom H.M. Ottenhoff (Leiden UMC), Nienke Paternotte (NWZ Alkmaar), Bart J.A. Rijnders (Erasmus MC), Anna H.E. Roukens (Leiden UMC), Esther Taks (Radboud), Janetta Top (UMC Utrecht), Karin M. Veerman (Radboud), Andreas Voss (CWZ Nijmegen), and Rob Willems (UMC Utrecht).

## Supporting information

CONSORT checklist

## Data Availability

All data produced in the present study are available upon reasonable request to the authors

## Acknowledgments

We thank the healthcare workers for their participation, Katina Kardaminidis for trial management, Frank Leus and Roxanne Schaakx for data management, and all other colleagues in the participating hospitals who implemented the BCG-Corona trial, and the sample collection and testing, under difficult circumstances.

## Author contributions

JW, TD, DB, FP, and MN designed and obtained funding for the study. JW supervised the data collection, analysis, and paper-writing. JC, TD, CG, TV, and AJ contributed to the data collection. GS, RB, and FK conducted the antibody testing and DB, LV and SJ provided additional immunological expertise. JC, TD, CG, and CW contributed to the data analyses. JC wrote the first draft of the manuscript. The additional members of the BCG-Corona study group contributed to the original BCG-Corona trial or other aspects of the ZonMw project (see Funding). All authors read and approved the final version. JW had full access to all the data in the study and takes responsibility for the integrity of the data and the accuracy of the data analysis.

