## Supplement for "BCG vaccination of healthcare workers does not reduce SARS-CoV-2 infections nor infection severity or duration: a randomised placebo-controlled trial"

### SUPPLEMENTARY METHODS

#### In-hospital sample collection and processing

Participants recruited in the three core hospitals (University Medical Center (UMC) Utrecht, Radboud UMC, and Leiden UMC) were invited to attend sampling visits in their own hospital at about 3 months (M3; June 2020) and 12 months (M12; April 2021) after study vaccination. They were asked to not eat or drink anything, and to not smoke, within the 30 minutes prior to the sampling in order to obtain high quality saliva samples (saliva data not included in this manuscript). Study nurses and physicians collected various blood samples via venepuncture and saliva samples using Denta sponge swabs. Participants donated about 10 ml blood in a serum (red cap) tube and in an EDTA plasma (purple cap) tube, and a subset of them donated an additional 18 ml blood in two 9 ml heparin serum (green cap) tubes for peripheral blood mononuclear cells (PBMC) isolation (plasma and PBMC data not included in this manuscript). The serum tubes were inverted 10 times immediately after collection, kept at room temperature, and taken to the laboratory within two hours for further processing in a level 2 biosafety laminar flow cabinet at room temperature. Serum tubes were allowed to coagulate for at least 30 min and centrifuged at 1200xg for 10 min. The supernatant was aliquoted into 3-4 2.0 ml screwcap tubes, which were stored frozen at -20 or -80 °C. A few weeks later, one serum tube per participant was transported on dry ice via courier to the Center for Immunology of Infections and Vaccines at the National Institute for Public Health and the Environment (RIVM in Dutch) in Bilthoven, the Netherlands, for antibody testing.

#### Fingerprick sampling at home

Participants recruited in the six other hospitals (Noordwest Ziekenhuis Alkmaar, Haga Ziekenhuis Den Haag, Canisius-Wilhelmina Ziekenhuis Nijmegen, Sint Maartenskliniek Nijmegen, Jeroen Bosch Ziekenhuis Den Bosch, and Erasmus Medisch Centrum Rotterdam), as well as participants from the three core hospitals who could not attend an in-hospital sampling visit, were asked via email if they were interested in receiving a home-sampling kit. Home sampling was coordinated by the UMC Utrecht team for all participating hospitals at about 6 months (M6) and 12 months (M12) after study vaccination. Each sampling round took several weeks to implement (M6: 09/10/2020 to 18/12/2020; M12: 14/04/2021 to 09/06/2021). Those wanting to participate were sent a sampling kit containing the following materials: a labelled 300 µl microtainer, 2 lancets, a gauze pad, a band-aid, a transparent sealable biospecimen bag, a labelled white medical post envelope (CoverMedPlus, Daklapack Europe) addressed to the RIVM with postage prepaid, and a participant instruction sheet. The participant was instructed to do the fingerprick, collect drops of blood in the microtainer, put the microtainer in the biospecimen bag together with the gauze, put the filled biospecimen bag in the white medical post envelope, and put the envelope in a regular mailbox that same day. At M6, participants received an email with a link to an online form to record their sample collection date, and at M12, this form also included questions about COVID-19-like symptoms, SARS-CoV-2 tests, and COVID-19 vaccinations between 27 March 2021 (diary app cessation) and the M12 sampling data. To maximise adherence, we contacted participants by email if the RIVM had not received the sample within a couple of weeks after mailing the kit.

**Antibody testing at the RIVM**

The RIVM team centrifuged samples immediately after receiving them, transferred the serum to a new tube, and froze the samples at -20°C until antibody testing. Serum samples were thawed on the day of testing. Total immunoglobulin G (IgG) antibody concentrations to SARS-CoV-2 Spike protein subunit S1 (anti-S1, all time points) and Nucleocapsid protein (anti-N, M12 only) were measured simultaneously using a bead-based assay, as previously described.[1]

IgG concentrations were calibrated against the international standard for human anti-SARS-CoV-2 immunoglobulin (20/136 NIBSC standard) and expressed as international units per mL (IU/mL).[2] The threshold for seropositivity was set at 10.1 IU/ml for anti-S1[3] and 14.3 IU/mL for anti-N antibodies.[4]

**Endpoint and follow-up time definitions**

The presence of infection episodes, and the duration and severity of each episode, was determined as described below by two independent researchers (JC and CG). They compared their individual coding and discussed discrepant cases. They also kept a list of special cases that were difficult to code. Persistent discrepant and special cases were discussed with the Principal Investigator (JW), who made the final coding decisions.

*Identifying infection episodes*

We assumed that few people had been exposed to SARS-CoV-2 prior to randomisation because the epidemic in the Netherlands had only just started and the first peak had not yet been reached. Indeed, only two participants reported to have had a positive test prior to randomisation, one of whom was included in our analysis population (see below). While SARS-CoV-2 testing was not widely available to the Dutch public at that time, it was available to HCWs. Blood samples were collected in two sampling rounds, dividing the follow-up time into two potential seroconversion periods. Period 1 is the period between study vaccination and the first sampling round, after about three months (M3) for the core hospitals and about 6 months (M6) for the other hospitals. Period 2 is the period between the first and the second sampling rounds, about 12 months (M12) after study vaccination for all hospitals. Infection episodes were categorised into the following categories, depending on available information for each participant for each study period:

<u>For period 1</u>:

1. Proven infection episode with seroconversion: Participant reported a positive test (with date) in the diary app, and had seroconverted (*developed anti-S1 antibodies*), in period 1.
2. Proven infection episode without seroconversion: Participant reported a positive test (with date) in the diary app in period 1, but there was no evidence of seroconversion (did not participate in the first sampling round or did not seroconvert).
3. Possible infection episode: Participant never reported a positive test in the diary app during period 1 but had seroconverted for anti-S1 antibodies in period 1. In some but not all cases, an episode start- and end-date could be estimated based on symptoms reported in the diary app (e.g. either one unique symptomatic period or multiple periods but all except one were accompanied by negative SARS-CoV-2 tests).

<u>For period 2</u>:

1. Proven infection episode with seroconversion: Participant reported a positive test (with date) in the diary app, and had seroconverted (*developed both anti-S1 and anti-N antibodies or anti-S1 if no COVID-vaccine reported*), in period 2.
2. Proven infection episode without seroconversion: Participant reported a positive test (with date) in the diary app in period 2, but there was no evidence of seroconversion (did not participate in the second sampling round or did not seroconvert).
3. Possible infection episode: Participant never reported a positive test in the diary app during

period 2 but had seroconverted in period 2. In some but not all cases, an episode start- and end-date could be estimated based on symptoms reported in the diary app (e.g. either one unique symptomatic period or multiple periods but all except one were accompanied by negative SARS-CoV-2 tests).

4. Inconclusive episode: Some participants tested positive for anti-N but not anti-S1 antibodies at the end of period 2. If they also did not report a positive SARS-CoV-2 test or a symptomatic episode, we considered these inconclusive episodes.

*Validating participant-reported positive SARS-CoV-2 tests*
In the first few months of the trial, the 9 participating hospitals asked their employees to test at their own hospital in case they had symptoms because public testing capacity was limited and self-tests were not available. Towards the end of the trial, public testing capacity increased but self-tests were still not available. We obtained test data from the UMC Utrecht Medical Microbiology laboratory for the BCG-Corona trial participants that were recruited in the UMC Utrecht. We were able to match the data reported by participants in the diary app to the laboratory data. A total of 1,164 tests were either reported by the 395 UMC Utrecht participants, by the lab, or both. This includes all testing episodes, not just the first one per participant. The agreement between participant reports and lab reports was high for positive tests but much lower for negative tests. We only used positive tests as endpoints in our analyses and the negative tests were therefore considered less important. Sixty-three positive tests were reported by both the participant and the lab, 17 by the participant but not the lab (these tests were likely done at a public testing site), and 26 by the lab but not the participant. In most of the latter cases (22/26), the participant was tested multiple times within a short time period (participants had to test negative before they could return to work) and only reported the first positive test of the series in the app. Therefore, only 4 positive tests should have been reported and were truly missed (two in each randomisation group); these 4 infections were not detected by serology either because no blood samples had been donated. In addition, 4 tests (three in the BCG and one in the placebo group) were reported positive by a participant but were negative according to the lab.

*Participant-reported positive SARS-CoV-2 tests in this paper versus the primary BCG-Corona paper*
The primary BCG-Corona paper analysed 210 participant-reported endpoints[15] whereas the current paper analysed 224. This difference can be explained by the slightly longer reporting period in the current paper (we added the period between the end of app completion on 27 March 2021 and the M12 sampling visits in April-June 2021) and the corrections that we made based on the laboratory data described above.

*Determining episode duration*
The duration of each infection episode was determined using symptoms and test results reported via the diary app. The acute infection episode duration was defined as the number of consecutive days during which the participant reported symptoms in the app, not including standalone loss of smell/taste or lingering symptoms (e.g. fatigue) after respiratory symptoms had ceased. The definition of "consecutive" was not strict: participants could have skipped reporting on one or two days within a consecutive period as long as symptoms were consistently reported before and after those days. Chronic symptoms that were reported throughout the study were ignored.

<u>Proven infection episodes with or without seroconversion:</u>
At least one positive test date is available for these infection episodes.

- *Start-date:* Defined as the date within the test window (14 days before to 14 days after the first positive test date) on which the first symptom was reported. If no symptoms were reported during the test window, the infection episode was considered asymptomatic and the start-date coded as not applicable.

- *End-date:* Defined as the date of the last reported symptom related to one episode. If the episode was asymptomatic, the end-date was coded as not applicable. If symptoms other than loss of smell/taste were ongoing on the last day of app entry, the end-date was coded as ongoing.

Possible infection episodes:
These infections were identified by seroconversion and did not have a positive test-date associated with them. If seroconversion occurred in period 2, all symptoms reported during period 1 were ignored.

- *Start-date:* All reported symptoms within the relevant study period(s) (period 1, 2, or both) were reviewed to determine if one or more acute symptoms episodes were present. If not, the infection episode responsible for the seroconversion was considered asymptomatic and the start-date coded as not applicable. If one or more symptoms episodes were identified, all episodes associated with a negative test were ignored. If only one symptoms episode remained, it was selected as the infection episode responsible for the seroconversion; the start-date was the day on which the first symptom of that episode was reported. When we could not be sure which symptoms episode was the infection episode, the start-date was coded as unknown.
- *End-date:* The date of the last reported symptom of the selected symptoms episode. If the episode was asymptomatic, or we could not be sure which symptoms episode was the infection episode, the end-dates were coded as not applicable or unknown, respectively.

*Categorising episode severity*
The following respiratory and non-respiratory symptoms could be reported in the diary app:

Respiratory symptoms (all reported on a scale of 0-5, with 0 meaning not present):

- Nose cold (*Neusverkouden*)
- Sore throat (*Keelpijn*)
- Cough (*Hoesten*)
- Dyspnoea/shortness of breath (*Kortademig*)
- Loss of smell/taste (*Reuk en/of smaakverlies*)

Non-respiratory symptoms (all reported on a scale of 0-5 except fever):

- Fever (*Koorts*), defined as a temperature of 38 °C or above
- Cold shivers (*Koude rillingen*)
- Muscle pain (*Spierpijn*)
- Fatigue (*Vermoeidheid*)
- Headache (*Hoofdpijn*)
- Diarrhoea (*Diarree*)

Categorisation of infection severity was not based on the World Health Organisation (WHO) common outcome measure set definitions because the WHO mild category is very broad, and the WHO moderate and severe categories were rare in our study population.[5] Instead, we based our definitions on the types and severity of reported symptoms (with a focus on respiratory symptoms and fever) as well as the episode duration, in our own dataset (Table S1). Chronic symptoms reported regularly throughout follow-up without a clear link to a testing date or an episode of respiratory symptoms/fever were ignored. This was, for example, often the case for headaches.

*Follow up time*
The number of follow-up days was calculated as the number of days between vaccination and the first infection episode (the positive test-date or the start-date of the symptomatic period if no test-date available; survival analyses) or the end of study (the M12 sampling date or the last day of app

completion if the participant did not provide a M12 sample; all other analyses).

*App completion percentage*
Participants were considered to have completed the diary app sufficiently if data were available for at least 80% of their follow-up days (Figure S1). The app completion percentage was the number of completed app entries divided by the expected number of app entries. Completed app entries are the number of days for which the participant completed the diary, regardless of whether or not the s/he formally withdrew from the study. The expected number of app entries is the number of days between the randomisation date and diary app cessation on 27 March 2021.

*Analysis population*
We started with the 1,511 randomised participants. We excluded 36 participants with inconclusive episodes but no participant-reported positive tests. A total of 206 participants had completed less than 80% of the diary app (one of whom also had an inconclusive episode), but 39 of them either self-reported a positive test or had evidence of seroconversion. In the case of these 39 participants, we can be sure that they experienced an endpoint. However, participants who did not have evidence of an endpoint and less than 80% diary app completion (N=166) could have experienced an endpoint without us finding out about it. We therefore excluded them. The total analysis population was therefore 1,309 participants and 298 events. Survival analyses included 1,252 participants and 241 events because 57 participants with an infection without start- and end-date could not be included.
We conducted several sensitivity analyses:

- We added back in the participants with less than 80% app completion and no evidence of infection, assuming that they did not have an infection.
- We added back in the participants with inconclusive episodes only, assuming that they were or were not true infection episodes.

**Statistical analysis**
The primary analyses consisted of comparing the cumulative incidence of SARS-CoV-2 infections, time to first SARS-CoV-2 infection, time to first mild-to-moderate, mild, and asymptomatic infection, and the episode duration between the BCG study group and the placebo groups.

Cumulative incidence was modelled in logistic regression models (all infections) or multinomial logistic regression (mild-to-moderate, mild, or asymptomatic infections) with randomisation group as the main predictor. Time-to-first-infection (any infection or mild-to-moderate, mild or asymptomatic infections) was modelled in Cox proportional hazards models with randomisation group as the main predictor. We started with unadjusted models and then adjusted for potential confounders. The following variables were considered as potential confounders a priori: recruitment site, enrolment week, age, sex, household size, smoking behaviour, several job-related characteristics (job function, percentage work hours with patient contact, scheduled to work on a COVID ward, and hospital department), past BCG vaccination, ever having tested positive for tuberculosis, current use of hypertension medication, current use of antidiabetic medication, history of pulmonary disease (hay fever, asthma, and any other pulmonary disease combined), and history of cardiovascular disease. We selected variables for inclusion in the final models using automatic backward selection, starting with the above-mentioned variables, and randomisation group forced into all models. The model selection settings using the Akaike Information Criterion based function ‘step’ in R were: lower scope=model with intervention as determinant (forced into model); upper scope=model with all covariates mentioned above; k=2.71; direction=backwards.

The assumptions of the Cox proportional hazards models were checked with scaled Schoenfeld residuals. We can assume proportional hazards because the global test for the relationship between residuals and time was non-significant, and the individual tests for all covariates in the multivariable model were also non-significant. We tested influential observations using dfbeta residuals. The results showed that none of the observations were very influential.

### SUPPLEMENTARY TABLES AND FIGURES

#### Table S1: Definitions of SARS-CoV-2 episode severity

| Severity | Definition |
|---|---|
| Hospitalisation | Hospitalisation due to COVID-19. This only applied to three cases and they were pooled with the mild-to-moderate category below. |
| Mild-to-moderate | Fever (temperature 38+) for more than one week AND/OR |
| | Dyspnea reaching level 4 or 5 for more than one week AND/OR |
| | Any other respiratory symptoms with at least one symptom other than loss of smell/taste reaching level 4 or 5 for more than one week AND/OR |
| | The total symptomatic episode lasted for more than 28 days[1] |
| Mild | Reported symptoms that were not chronic throughout the study but did not reach the level of mild-to-moderate as described above. |
| Asymptomatic | Infection episode detected via SARS-CoV-2 diagnostic testing or serology but no symptoms reported during the relevant time period. |
| Unknown | It is impossible to draw conclusions from the available data. |
| Long-COVID | Continuing to report symptoms for at least 60 consecutive days after the end of the acute infection episode. If loss of smell-taste was the only lingering symptom, this was coded as long-term loss of smell/taste and not long COVID. |
| Long-term loss of smell/taste | Lingering loss of smell/taste for at least 60 consecutive days after the end of acute infection episode. |

1. We made an exception for two participants who had a nose cold at level 1-2 for more than 4 weeks and nothing else.

**Figure S1: Completeness of smartphone diary application entries by A) all randomised participants (N=1511), and B) all participants that experienced an event (N=298)**

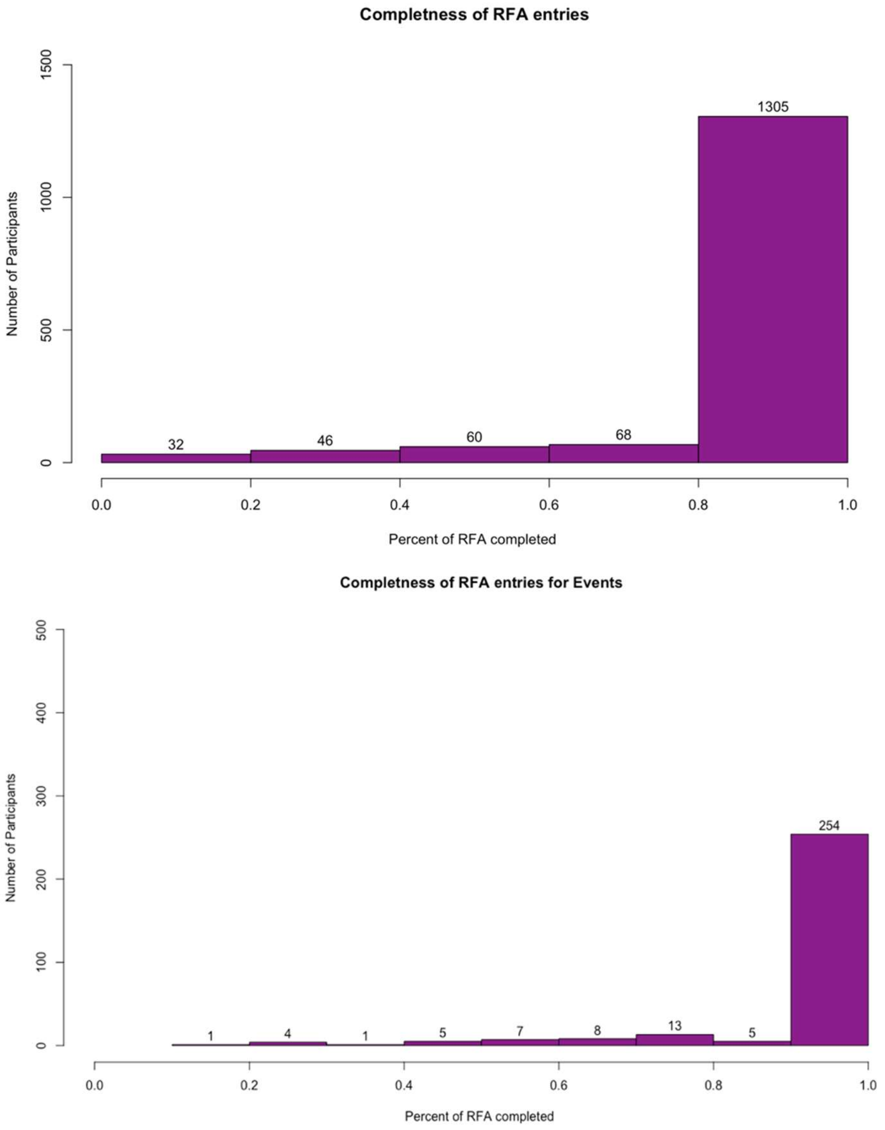


A) Completeness of diary entries for the entire randomised population (N=1511); completion percentages were not significantly different between randomisation groups (p=0.283).

B) Completeness of diary entries for those that experienced an event (N=298). The 39 participants who did not complete at least 80% of expected diary entries were included in the analysis population because we did have clear evidence that they had an infection.

**Table S2: Baseline characteristics of the randomised population**

| Cells contain n (% of N) unless stated otherwise | | BCG (N=753) | Placebo (N=758) | Total (N=1,511) | p[1] |
|---|---|---|---|---|---|
| **Recruitment site** | Radboud UMC | 209 (27.8) | 210 (27.7) | 419 (27.7) | 1.000 |
| | UMC Utrecht | 193 (25.6) | 192 (25.3) | 385 (25.5) | |
| | Noordwest ZH Alkmaar | 158 (21.0) | 158 (20.8) | 316 (20.9) | |
| | Haga ZH Den Haag | 50 (6.6) | 52 (6.9) | 102 (6.8) | |
| | Canisius-Wilhelmina ZH Nijmegen | 38 (5.0) | 37 (4.9) | 75 (5.0) | |
| | Sint Maartenskliniek Nijmegen | 31 (4.1) | 34 (4.5) | 65 (4.3) | |
| | Leiden UMC | 28 (3.7) | 28 (3.7) | 56 (3.7) | |
| | Jeroen Bosch ZH Den Bosch | 25 (3.3) | 26 (3.4) | 51 (3.4) | |
| | Erasmus UMC | 21 (2.8) | 21 (2.8) | 42 (2.8) | |
| **Age in years, mean (SD)[2]** | | 41.31 (12.63) | 42.76 (12.73) | 42.04 (12.70) | **0.026** |
| **Female sex** | | 572 (76.0) | 550 (72.6) | 1122 (74.3) | 0.146 |
| **# Additional household members, mean (SD)** | | 2.00 (1.53) | 1.85 (1.41) | 1.93 (1.47) | 0.056 |
| **Smoking status** | Current | 63 (8.4) | 60 (7.9) | 123 (8.1) | 0.407 |
| | Former | 228 (30.3) | 208 (27.4) | 436 (28.9) | |
| | Never | 462 (61.4) | 490 (64.6) | 952 (63.0) | |
| **Hospital department** | Urgent care | 40 (5.3) | 47 (6.2) | 87 (5.8) | 0.529 |
| | Internal medicine[3] | 127 (16.9) | 109 (14.4) | 236 (15.6) | |
| | Intensive/medium care | 67 (8.9) | 72 (9.5) | 139 (9.2) | |
| | Other | 519 (68.9) | 530 (69.9) | 1049 (69.4) | |
| **Job function** | Doctor | 172 (22.8) | 185 (24.4) | 357 (23.6) | 0.643 |
| | Nurse | 363 (48.2) | 367 (48.4) | 730 (48.3) | |
| | Paramedic | 123 (16.3) | 107 (14.1) | 230 (15.2) | |
| | Support personnel | 95 (12.6) | 99 (13.1) | 194 (12.8) | |
| **Scheduled to work on COVID-ward** | No | 226 (30.0) | 219 (28.9) | 445 (29.5) | 0.478 |
| | Yes | 481 (63.9) | 481 (63.5) | 962 (63.7) | |
| | Unknown | 46 (6.1) | 58 (7.7) | 104 (6.9) | |
| **% work hours with patient contact** | 0-25 | 111 (14.7) | 127 (16.8) | 238 (15.8) | 0.163 |
| | 26-50 | 120 (15.9) | 93 (12.3) | 213 (14.1) | |
| | 51-75 | 131 (17.4) | 126 (16.6) | 257 (17.0) | |
| | 75+ | 391 (51.9) | 412 (54.4) | 803 (53.1) | |
| **History of BCG vaccination** | | 129 (17.1) | 127 (16.8) | 256 (16.9) | 0.899 |
| **Past tuberculosis test results[4]** | | | | | |
| Tested negative | | 504 (66.9) | 512 (67.5) | 1016 (67.2) | |
| Tested positive (either or both) | | 58 (7.7) | 78 (10.3) | 136 (9.0) | **0.030** |
| Never tested | | 181 (24.0) | 166 (21.9) | 347 (23.0) | |
| Unknown (both) | | 10 (1.3) | 2 (0.3) | 12 (0.8) | |
| **Respiratory infection in winter 2019-2020** | | | | | |
| No | | 548 (72.8) | 542 (71.5) | 1090 (72.1) | 0.270 |
| Yes, with fever | | 69 (9.2) | 58 (7.7) | 127 (8.4) | |
| Yes, no fever | | 136 (18.1) | 158 (20.8) | 294 (19.5) | |
| **Influenza vaccination in winter 2020-2021** | | | | | 0.610 |
| Yes | | 342 (45.4) | 355 (46.8) | 697 (46.1) | |
| No | | 222 (29.5) | 206 (27.2) | 428 (28.3) | |
| Missing | | 189 (25.1) | 197 (26.0) | 386 (25.5) | |
| **Any other vaccination in past year[5]** | | 85 (11.3) | 77 (10.2) | 162 (10.7) | 0.531 |
| **Current use of anti-hypertensive medication** | | 49 (6.5) | 50 (6.6) | 99 (6.6) | 1.000 |
| **History of cardiovascular disease** | | 15 (2.0) | 19 (2.5) | 34 (2.3) | 0.616 |
| **Current use of anti-diabetic medication** | | 3 (0.4) | 6 (0.8) | 9 (0.6) | 0.510 |
| **History of asthma** | | 54 (7.2) | 47 (6.2) | 101 (6.7) | 0.514 |
| **History of hay fever** | | 229 (30.4) | 212 (28.0) | 441 (29.2) | 0.323 |
| **History of other pulmonary disease** | | 14 (1.9) | 18 (2.4) | 32 (2.1) | 0.605 |

| | | | | |
|---|---|---|---|---|
| **Any lung disease (previous three combined)[6]** | 254 (33.7) | 243 (32.1) | 497 (32.9) | 0.452 |
| **Positive SARS-CoV-2 test prior to baseline** | 1 (0.1) | 1 (0.11) | 2 (0.1) | 1.000 |
| **At least one dose COVID-19 vaccine during FU[7]** | 362 (43.3) | 335 (44.2) | 661 (43.7) | 0.736 |
| **COVID-19 vaccine type[7]** | | | | |
| Comirnaty | 203 (27.0) | 198 (26.1) | 401 (26.5) | 0.815 |
| Spikevax | 69 (9.2) | 80 (10.6) | 149 (9.9) | |
| Vaxzevria | 44 (5.8) | 44 (5.8) | 88 (5.8) | |
| Jcovden | 7 (0.9) | 6 (0.8) | 13 (0.9) | |
| CureVac | 0 (0.0) | 1 (0.1) | 1 (0.1) | |
| Unknown | 3 (0.4) | 6 (0.8) | 9 (0.6) | |
| None | 427 (56.7) | 423 (55.8) | 850 (56.3) | |

Abbreviations: FU=follow-up; SD=standard deviation; UMC=University Medical Center; ZH=ziekenhuis (hospital).

1. Chi-squared tests for categorical variables and Wilcoxon rank sum test for continuous variables.
2. The 1.45 years difference in mean age is statistically significant but we believe that it is not relevant in this context.
3. Internal medicine includes the pulmonology and infectious disease departments.
4. Tuberculosis tests include the Mantoux and/or TB QuantiFERON tests. The statistical difference between the BCG and placebo groups is for the unknown category only.
5. The following other vaccinations were reported: DTaP-IPV, hepatitis A, hepatitis B, yellow fever, typhoid, rabies, mumps-measles-rubella, meningococcal, pneumococcal, *Haemophilus influenzae* type B, Ebola, tick-borne encephalitis, human papillomavirus, and unknown.
6. “Any lung disease” includes asthma, hay fever, and any other pulmonary disease.
7. In the Netherlands, the SARS-CoV-2 vaccines available during the study period were Comirnaty (Pfizer/BioNTech, New York, NY, USA), Spikevax (Moderna Biotech, Cambridge, MA, USA), Vaxzevria (AstraZeneca AB, Sodertalje, Sweden), and Jcovden (Janssen Vaccines, Leiden, Netherlands). In addition, one participant received an experimental vaccine by CureVac N.V. in a clinical trial setting. This vaccine was never marketed due to insufficient efficacy.

**Figure S2: SARS-CoV-2 infections over calendar time in the study population for all infections (A), by infection severity (B), by recruitment site (C), and in the Netherlands as a whole (D)**

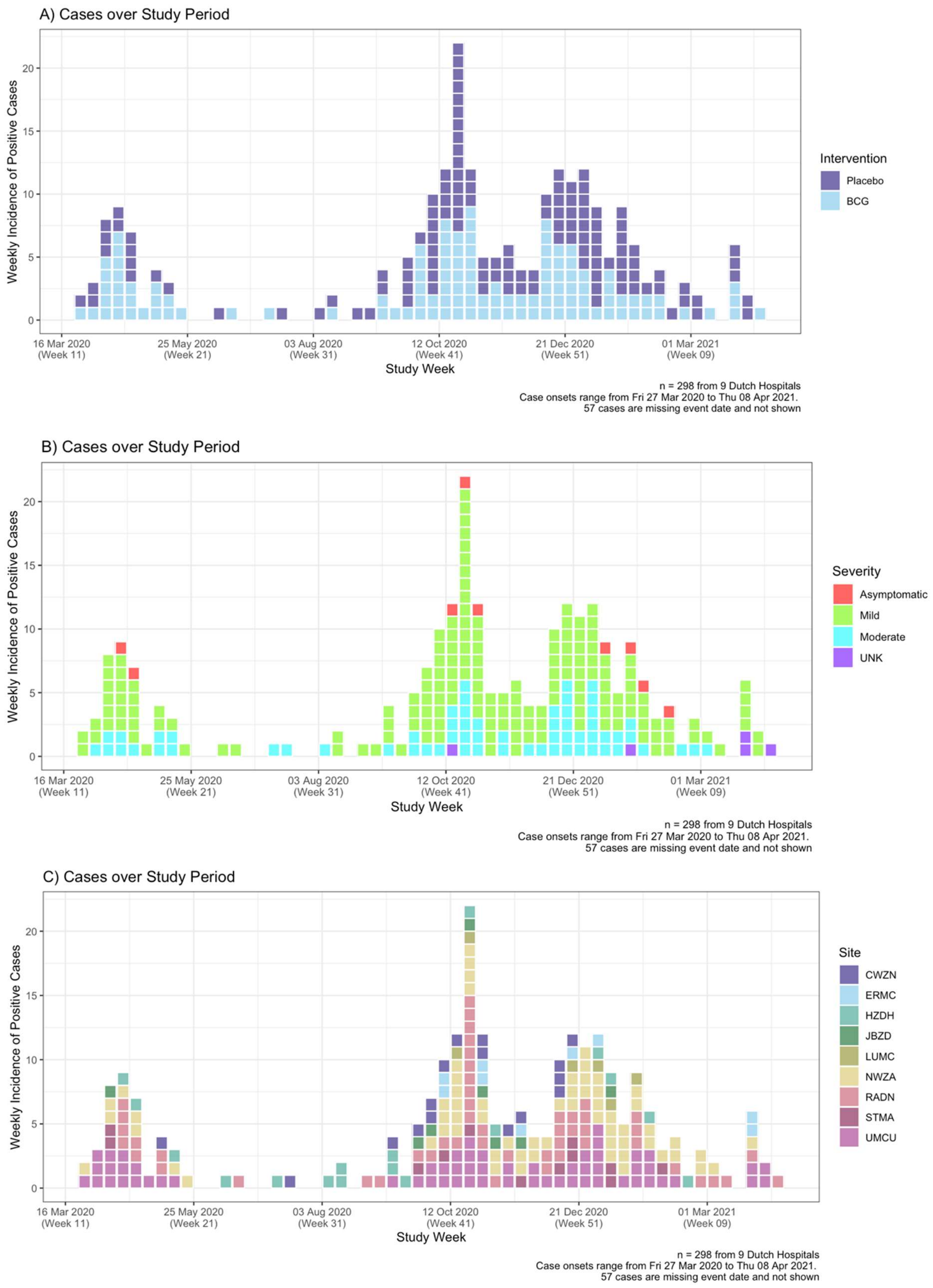

D)

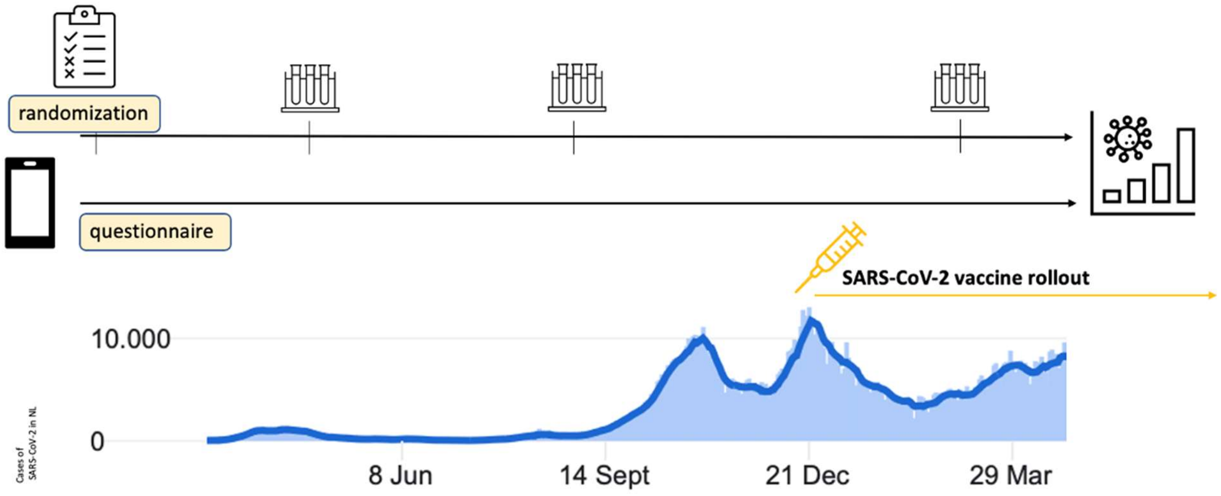


**Panel D:** Incidence of SARS-CoV-2 cases in the Netherlands during the study follow-up period. SARS-CoV-2 infections over calendar time in the study population mirrored epidemic waves in the Netherlands except for the March-May 2020 wave. Infections in the general public were underdetected during that wave because public testing was not yet widely available. In contrast, healthcare workers could get tested during that wave in the hospitals where they worked.

Data sources:

1) The Corona dashboard of the Dutch National Institute for Health and Environment (RIVM): https://coronadashboard.rijksoverheid.nl/
2) COVID-19 Data Repository by the Center for Systems Science and Engineering at Johns Hopkins University (Dong E, Du H, Gardner L. An interactive web-based dashboard to track COVID-19 in real time. *Lancet Inf Dis* 2020; 20(5):533-534. doi: 10.1016/S1473-3099(20)30120-1).

### Table S3: Additional outcomes during follow-up in the analysis population[1]

| N=participants | BCG (N=665) | Placebo (N=644) | Total (N=1,309) | p[2] |
|---|---|---|---|---|
| Peron-years at risk | 597.08 | 585.49 | 1,182.57 | |
| Incidence rate<br>Incidence rate ratio (95% CI) | 0.25 | 0.26 | <br>0.95 (0.76, 1.21) | 0.732 |
| Cumulative incidence | 0.22 | 0.23 | 0.23 | 0.608 |
| Anti-S1 antibodies present in round 2[3,4], n (%) | 271 (49.2) | 275 (49.8) | 546 (49.5) | 0.880 |
| Anti-N antibodies present in round 2[4], n (%) | 75 (13.6) | 88 (16.0) | 163 (14.8) | 0.315 |
| **N=episodes** | **BCG (N=147)** | **Placebo (N=151)** | **Total (N=298)** | **p[2]** |
| Period that episode occurred in, n (%): | | | | |
| Period 1 | 57 (38.8) | 34 (22.5) | 91 (30.5) | **0.007** |
| Period 2 (or period 1 if no round 1 done) | 89 (60.5) | 114 (75.5) | 203 (68.1) | |
| Unknown | 1 (0.7) | 3 (2.0) | 4 (1.3) | |
| Seroconversion status after episode, n (%): | | | | |
| Did not seroconvert | 8 (5.4) | 16 (10.6) | 24 (8.1) | **0.002** |
| Episode in period 1, S+ in round 1 | 46 (31.3) | 23 (15.2) | 69 (23.2) | |
| Episode in period (1+)2, S+N+ in round 2 | 57 (38.8) | 69 (45.7) | 126 (42.3) | |
| Episode in period (1+)2, S+N- in round 2 | 9 (6.1) | 21 (13.9) | 30 (10.1) | |
| Episode in period (1+)2, S-N+ in round 2 | 0 (0.0) | 2 (1.3) | 2 (0.7) | |
| Unknown (no serology done or unknown period) | 27 (18.4) | 20 (13.2) | 47 (15.8) | |
| Fever during episode, n (%) | 50 (34.0) | 46 (30.5) | 96 (32.2) | 0.803 |
| Unknown | 3 (2.0) | 3 (2.0) | 6 (2.0) | |
| Mean fever duration in days (SD) | 1.86 (3.30) | 2.20 (4.62) | 2.03 (4.02) | 0.477 |
| Dyspnea level,[5] n (%): | | | | |
| 0 | 101 (68.7) | 107 (70.9) | 208 (69.8) | 0.823 |
| 1-3 | 28 (19.0) | 29 (19.2) | 57 (19.1) | |
| 4-5 | 16 (10.9) | 12 (7.9) | 28 (9.4) | |
| Unknown/missing | 2 (1.4) | 3 (2.0) | 5 (1.7) | |
| Mean dyspnoea duration in days (SD) | 2.86 (7.99) | 3.40 (10.65) | 3.13 (9.43) | 0.627 |
| Other respiratory symptoms level,[5,6] n (%): | | | | |
| 0 | 27 (18.4) | 29 (19.2) | 56 (18.8) | 0.987 |
| 1-3 | 59 (40.1) | 62 (41.1) | 121 (40.6) | |
| 4-5 | 49 (33.3) | 49 (32.5) | 98 (32.9) | |
| Unknown/missing | 12 (8.2) | 11 (7.3) | 23 (7.7) | |
| Mean respiratory symptoms duration in days (SD) | 8.70 (8.92) | 9.24 (9.92) | 8.98 (9.44) | 0.644 |
| Non-respiratory symptoms level,[6] n (%): | | | | |
| 0 | 32 (21.8) | 32 (21.2) | 64 (21.5) | 0.944 |
| 1-3 | 31 (21.1) | 29 (19.2) | 60 (20.1) | |
| 4-5 | 73 (49.7) | 80 (53.0) | 153 (51.3) | |
| Unknown/missing | 11 (7.5) | 10 (6.6) | 21 (7.0) | |
| Total symptoms duration in weeks, n (%) | | | | |
| 0 days (asymptomatic) | 23 (15.6) | 19 (12.6) | 42 (14.1) | 0.472 |
| Up to 1 week | 16 (10.9) | 28 (18.5) | 44 (15.1) | |
| 1-2 weeks | 35 (23.8) | 29 (19.2) | 64 (21.5) | |
| 2-3 weeks | 24 (16.3) | 21 (13.9) | 45 (15.1) | |
| 3-4 weeks | 9 (6.1) | 12 (7.9) | 21 (7.0) | |
| More than 4 weeks | 22 (15.0) | 27 (17.9) | 49 (16.4) | |
| Unknown (including ongoing at end of study) | 18 (12.2) | 15 (9.9) | 33 (11.1) | |

Abbreviations: SD=standard deviation.

1. The main outcomes are shown in Table 2 of the manuscript.
2. Chi-square tests for categorical variables and Wilcoxon rank sum test for means.
3. Includes the participants who had received at least one dose of a COVID-19 vaccine. They were equally distributed among the randomisation groups (Table 1 of the manuscript).

4. The denominators are 551 (665 minus 114 who did not provide a M12 sample) in the BCG group, 552 (644 minus 92) in the placebo group, and 1,103 in the total analysis population.
5. Highest level reported by the participant during the entire infection episode.
6. Does not include dyspnoea (see separate mean duration for dyspnoea) or loss of smell and/or taste.

### Figure S3: Infection episode duration distributions by randomisation group

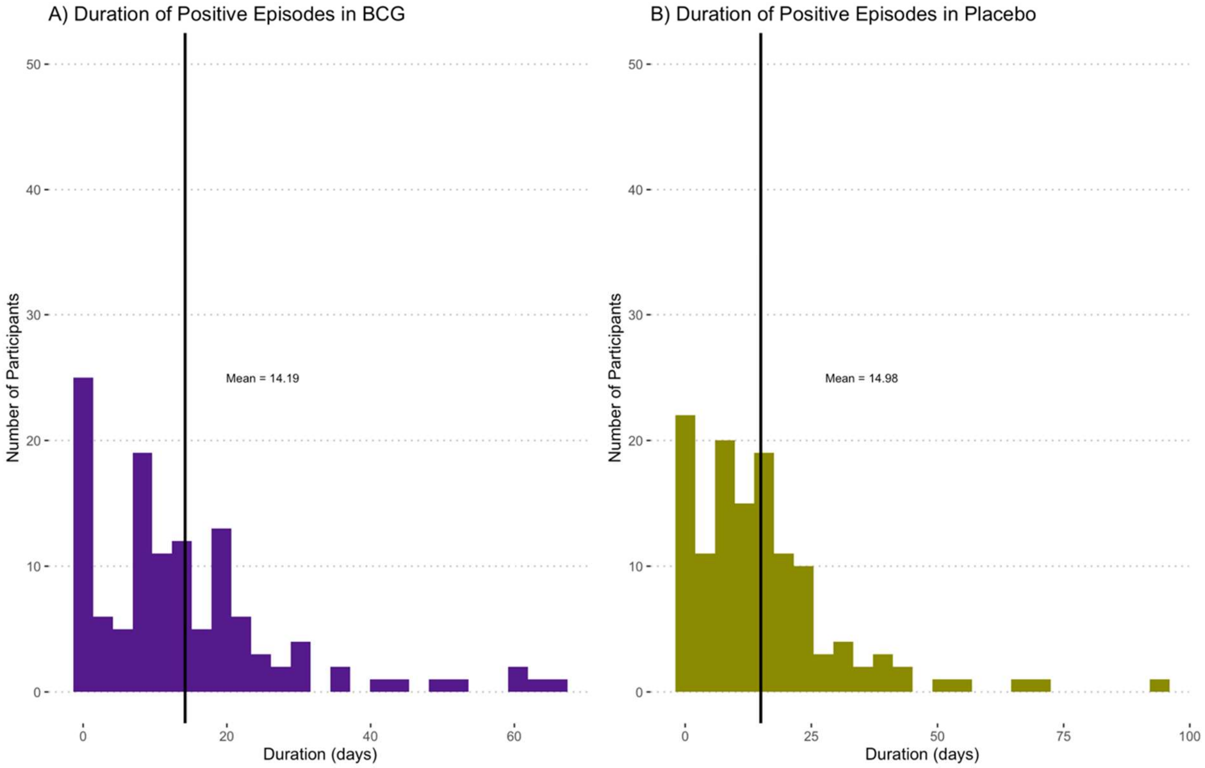


1. N=248 infection episodes: 49 episodes with unknown or ongoing duration at the end of study, and one outlier with duration of 398 are not shown (p=0.724).
2. A non-parametric test (Wilcoxon) was used to compare the mean duration between the groups. For the calculation of acute episode duration cases of long COVID-19 and episodes with unknow or ongoing duration were removed; N=189 and the difference in mean duration remained non-significant (0.890) (Table 2)

### Table S4: Univariable regression models with SARS-CoV-2 infection as outcome

| Covariate | Logistic regression | | Cox regression | |
|---|---|---|---|---|
| | OR (95% CI) | p | HR (95% CI) | p |
| Age in years | 0.99 (0.98, 1.00) | **0.005** | 0.99 (0.98, 1.00) | **0.005** |
| Female sex | 1.34 (0.99, 1.84) | 0.065 | 1.32 (0.97, 1.80) | 0.075 |
| Additional household member | 1.06 (0.97, 1.16) | 0.217 | 1.08 (0.99, 1.17) | 0.101 |
| Smoking status | | | | |
| Current | -- | -- | -- | -- |
| Former | 1.19 (0.71, 2.07) | 0.531 | 1.27 (0.75, 2.13) | 0.374 |
| Never | 0.98 (0.59, 1.66) | 0.924 | 0.99 (0.60, 1.63) | 0.953 |
| Hospital department | | | | |
| Urgent care | -- | -- | -- | -- |
| Internal medicine[1] | 1.21 (0.68, 2.20) | 0.520 | 1.18 (0.68, 2.03) | 0.557 |
| Intensive/Medium care | 0.60 (0.31, 1.18) | 0.135 | 0.45 (0.22, 0.91) | **0.027** |
| Other | 0.68 (0.40, 1.17) | 0.146 | 0.71 (0.43, 1.17) | 0.176 |
| Job function | | | | |
| Doctor | -- | -- | -- | -- |
| Nurse | 1.95 (1.38, 2.80) | **<0.001** | 1.81 (1.27, 2.58) | **0.001** |
| Paramedic | 1.29 (0.81, 2.04) | 0.281 | 1.26 (0.79, 2.00) | 0.335 |
| Support staff | 1.18 (0.71, 1.93) | 0.522 | 1.21 (0.74, 1.99) | 0.452 |
| Scheduled to work on COVID-ward | | | | |
| No | -- | -- | -- | -- |
| Yes | 1.98 (1.45, 2.74) | **<0.001** | 1.75 (1.27, 2.39) | **<0.001** |
| Unknown | 1.55 (0.85, 2.73) | 0.139 | 1.47 (0.84, 2.58) | 0.179 |
| % work hours with patient contact: | | | | |
| 0-25 | -- | -- | -- | -- |
| 26-50 | 1.68 (0.98, 2.93) | 0.061 | 1.69 (0.97, 2.92) | 0.063 |
| 51-75 | 1.67 (0.99, 2.87) | 0.058 | 1.68 (0.98, 2.88) | 0.059 |
| 75+ | 2.68 (1.75, 4.25) | **<0.001** | 2.37 (1.50, 3.75) | **<0.001** |
| History of BCG vaccination | 1.14 (0.81, 1.59) | 0.428 | 1.12 (0.81, 1.55) | 0.496 |
| Past tuberculosis test results[2] | | | | |
| Negative result | -- | -- | -- | -- |
| Positive result | 1.18 (0.75, 1.82) | 0.454 | 1.10 (0.72, 1.69) | 0.697 |
| Never Done | 1.27 (0.93, 1.72) | 0.133 | 1.06 (0.78, 1.45) | 0.697 |
| Unknown | 0.73 (0.11, 2.78) | 0.681 | 0.90 (0.22, 3.64) | 0.886 |
| Current use of anti-hypertensive medication | 1.50 (0.92, 2.38) | 0.092 | 1.53 (1.00, 2.33) | **0.049** |
| History of cardiovascular disease | 1.24 (0.51, 2.71) | 0.607 | 1.13 (0.50, 2.53) | 0.775 |
| Current use of anti-diabetic medication | 1.36 (0.19, 6.34) | 0.714 | 1.50 (0.37, 6.04) | 0.567 |
| History of asthma | 1.09 (0.65, 1.76) | 0.739 | 1.21 (0.77, 1.91) | 0.414 |
| History of hay fever | 1.19 (0.89, 1.57) | 0.231 | 1.16 (0.89, 1.53) | 0.272 |
| History of other pulmonary disease[3] | 1.54 (0.66, 3.33) | 0.286 | 1.43 (0.67, 3.03) | 0.354 |
| Any Lung Disease[4] | 1.21 (0.92, 1.58) | 0.166 | 1.19 (0.92, 1.55) | 0.188 |
| BCG Vaccination[5] | | | | |
| Never | -- | -- | -- | -- |
| Past | 0.86 (0.51, 1.40) | 0.563 | 0.89 (0.55, 1.45) | 0.636 |
| Trial | 0.84 (0.63, 1.12) | 0.230 | 0.85 (0.64, 1.12) | 0.249 |
| Past and Trial | 1.24 (0.78, 1.92) | 0.352 | 1.17 (0.76, 1.80) | 0.469 |

Abbreviations: OR=odds ratio; HR=hazards ratio; 95% CI= 95% confidence interval.

1. Internal medicine includes the pulmonology and infectious disease departments
2. Tuberculosis tests include the Mantoux and/or TB QuantiFERON tests. The statistical difference between the BCG and placebo groups is for the unknown category only.
3. This does not include history of asthma or hay fever.
4. “Any lung disease” includes asthma, hay fever, and any other pulmonary disease.
5. Summary variable for participants that received a BCG vaccination is the past, in the trial, both, or never.

**Table S5: Multinomial logistic regression models with infection severity as the outcome[1]**

| Covariates | Outcome categories | OR (95% CI) | p |
|---|---|---|---|
| BCG versus placebo | No infection | -- | -- |
| | Asymptomatic | 1.13 (0.60, 2.11) | 0.711 |
| | Mild | 0.87 (0.62, 1.22) | 0.417 |
| | Mild-to-moderate | 0.71 (0.42, 1.21) | 0.202 |
| Age per year | No infection | -- | -- |
| | Asymptomatic | 0.99 (0.96, 1.02) | 0.391 |
| | Mild | 0.97 (0.96, 0.99) | <0.**001** |
| | Mild-to-moderate | 1.00 (0.98, 1.03) | 0.734 |
| Working in COVID-ward: yes vs. no | No infection | -- | -- |
| | Asymptomatic | 1.73 (0.73, 4.12) | 0.216 |
| | Mild | 1.34 (0.88, 2.05) | 0.177 |
| | Mild-to-moderate | 2.97 (1.36, 6.04) | **0.006** |
| Working in COVID-ward: unknown vs. no | No infection | -- | -- |
| | Asymptomatic | 1.73 (0.43, 6.88) | 0.440 |
| | Mild | 1.30 (0.63, 2.67) | 0.479 |
| | Mild-to-moderate | 1.36 (0.35, 5.23) | 0.655 |
| Hospital department: internal medicine vs. urgent care | No infection | -- | -- |
| | Asymptomatic | 0.92 (0.26, 3.24) | 0.898 |
| | Mild | 0.98 (0.49, 1.95) | 0.961 |
| | Mild-to-moderate | 8.47 (1.07, 67.3) | **0.043** |
| Hospital department: intensive/medium care vs. urgent care | No infection | -- | -- |
| | Asymptomatic | 0.87 (0.24, 3.13) | 0.823 |
| | Mild | 0.19 (0.07, 0.53) | **0.001** |
| | Mild-to-moderate | 3.78 (0.45, 31.9) | 0.222 |
| Hospital department: other vs. urgent care | No infection | -- | -- |
| | Asymptomatic | 0.56 (0.18, 1.77) | 0.326 |
| | Mild | 0.64 (0.34, 1.19) | 0.156 |
| | Mild-to-moderate | 4.91 (0.65, 37.2) | 0.123 |
| Hospital function: nurse vs. doctor | No infection | -- | -- |
| | Asymptomatic | 1.94 (0.82, 4.63) | 0.132 |
| | Mild | 1.87 (1.19, 2.94) | **0.007** |
| | Mild-to-moderate | 3.87 (1.60, 9.36) | **0.003** |
| Hospital function: paramedic vs. doctor | No infection | -- | -- |
| | Asymptomatic | 1.08 (0.30, 3.85) | 0.908 |
| | Mild | 1.59 (0.89, 2.83) | 0.117 |
| | Mild-to-moderate | 1.69 (0.52, 5.44) | 0.380 |
| Hospital function: support staff vs. doctor | No infection | -- | -- |
| | Asymptomatic | 1.90 (0.56, 6.42) | 0.303 |
| | Mild | 1.07 (0.53, 2.15) | 0.852 |
| | Mild-to-moderate | 3.27 (1.06, 10.1) | **0.039** |
| Past BCG vaccination | No infection | -- | -- |
| | Asymptomatic | 1.83 (0.81, 4.12) | 0.146 |
| | Mild | 1.85 (1.16, 2.95) | **0.009** |
| | Mild-to-moderate | 1.15 (0.55, 2.41) | 0.706 |
| Current use of hypertension medication | No infection | -- | -- |
| | Asymptomatic | 1.24 (0.35, 4.36) | 0.736 |
| | Mild | 2.05 (1.10, 3.84) | **0.024** |
| | Mild-to-moderate | 2.05 (0.85, 4.94) | 0.108 |

Abbreviations: OR=odds ratio; 95% CI= 95% confidence interval.

1. N=1,309 participants and 298 endpoints with cumulative asymptomatic, mild, or mild-to-moderate SARS-CoV-2 infections as endpoints. In addition to the variables listed in this table, the following variables were considered for inclusion in the model: recruitment site, enrolment week, sex, additional household members, percentage of work hours in contact with patients, ever having tested positive for tuberculosis, smoking status, current use of antidiabetic medication, history of pulmonary disease, and history of cardiovascular disease.

**Table S6: Sensitivity analysis adding back in the participants with less than 80% diary app completion and no evidence of infection, assuming that they never had an infection**

### A) Logistic regression models (cumulative incidence)

| BCG vs placebo | | Analysis population[1]<br><br>N=1,309<br>298 episodes | Sensitivity analysis population[2]<br>N=1,475<br>298 episodes |
|---|---|---|---|
| | | **OR (95% CI); p** | **OR (95% CI); p** |
| **Unadjusted** | | | |
| All episodes | | 0.93 (0.72, 1.20); 0.563 | 0.98 (0.76, 1.27); 0.887 |
| Participant-reported episodes only | | 0.89 (0.66, 1.18); 0.413 | 0.94 (0.71, 1.25); 0.666 |
| Asymptomatic episodes only | | 1.15 (0.62, 2.16); 0.654 | 1.22 (0.66,2.29); 0.527 |
| Mild episodes only | | 0.94 (0.68, 1.31); 0.714 | 1.00 (0.72, 1.38); 0.983 |
| Mild-to-moderate episodes only | | 0.73 (0.43, 1.23); 0.241 | 0.78 (0.46, 1.30); 0.339 |
| **Adjusted for site and enrolment week** | | | |
| All episodes | | 0.93 (0.72, 1.20); 0.577 | 0.98 (0.76, 1.26); 0.869 |
| Participant-reported episodes only | | 0.88 (0.66, 1.18); 0.395 | 0.93 (0.69,1.24); 0.606 |
| **Adjusted for age and sex** | | | |
| All episodes | | 0.90 (0.70, 1.17); 0.443 | 0.96 (0.74, 1.24); 0.762 |
| Participant-reported episodes only | | 0.86 (0.64, 1.15); 0.304 | 0.91 (0.69, 1.22); 0.539 |
| **Multivariable (backward selection)[3]** | | | |
| All episodes | | 0.85 (0.65, 1.12); 0.249 | 0.92 (0.71, 1.20); 0.541 |
| Participant-reported episodes only | | 0.81 (0.59, 1.09); 0.161 | 0.87 (0.65, 1.17); 0.353 |
| **Multinomial** | | | |
| Unadjusted<br>All episodes | Asymptomatic | 1.15 (0.62, 2.14); 0.654 | 1.22 (0.66, 2.27); 0.526 |
| | Mild | 0.94 (0.68, 1.31); 0.714 | 1.00 (0.72, 1.38); 0.983 |
| | Mild-to-moderate | 0.73 (0.44, 1.23); 0.241 | 0.78 (0.46, 1.30); 0.339 |
| Unadjusted<br>Participant-reported only | Asymptomatic | 0.95 (0.27, 3.31); 0.937 | 1.01 (0.29, 3.52); 0.984 |
| | Mild | 0.94 (0.67, 1.32); 0.720 | 1.00 (0.71, 1.40); 0.979 |
| | Mild-to-moderate | 0.73 (0.43, 1.23); 0.236 | 0.77 (0.46, 1.30); 0.331 |
| Adjusted for site, enrolment week<br>All episodes | Asymptomatic | 1.15 (0.62, 2.14); 0.663 | 1.23 (0.66, 2.28); 0.517 |
| | Mild | 0.95 (0.68, 1.32); 0.747 | 1.00 (0.72, 1.38); 0.975 |
| | Mild-to-moderate | 0.73 (0.44, 1.23); 0.243 | 0.77 (0.46, 1.30); 0.329 |
| Adjusted for age and sex<br>All episodes | Asymptomatic | 1.13 (0.61, 2.10); 0.699 | 1.20 (0.65, 2.23); 0.560 |
| | Mild | 0.91 (0.65, 1.27); 0.573 | 0.97 (0.70, 1.34); 0.850 |
| | Mild-to-moderate | 0.73 (0.43, 1.22); 0.230 | 0.77 (0.46, 1.30); 0.329 |
| Multivariable (backward selection)[4] | Asymptomatic | 1.13 (0.60, 2.11); 0.711 | 1.20 (0.64, 2.25); 0.560 |
| | Mild | 0.87 (0.62, 1.22); 0.417 | 0.94 (0.67, 1.31); 0.699 |
| | Mild-to-moderate | 0.71 (0.41, 1.21); 0.202 | 0.76 (0.45, 1.28); 0.304 |

Abbreviations: OR=odds ratio; 95% CI= 95% confidence interval.

1. Participant with no evidence of an infection episode who completed less than 80% of the expected diary app entries were excluded because we could not be sure that they never had an infection. Participants with inconclusive infection episodes (N=36) were also removed.
2. All participants were included regardless of diary app completion, except participants with inconclusive episodes (N=36). The sensitivity analysis assumed that participant with no evidence of an infection episode who completed less than 80% of the expected diary app entries did in fact never have an infection.
3. Covariates considered as potential confounders are shown in Table S4. Covariates retained in the model were: age in years, additional number of household members, function, % work hours with patient contact, hospital department, expected to work in COVID-ward, past history of BCG vaccination, and current use of hypertension medication.
4. Covariates considered as potential confounders are shown in Table S4. Covariates retained in the model were: age in years, hospital function and department, expected to work in COVID-ward, history of BCG vaccination, and current use of hypertension medication.

### B) Cox regression models (time to first infection)

| BCG vs. placebo | Analysis population[1]<br>N=1,252<br>241 episodes | Sensitivity analysis population[2]<br>N=1,403<br>241 episodes |
|---|---|---|
| | **HR (95% CI); p** | **HR (95% CI); p** |
| **Unadjusted** | | |
| All episodes | 0.92 (0.72, 1.19); 0.521 | 0.92 (0.72, 1.19); 0.532 |
| Participant-reported episodes only | 0.91 (0.70, 1.19); 0.493 | 0.91 (0.70, 1.19); 0.496 |
| Asymptomatic episodes only | 1.19 (0.32, 4.44); 0.793 | 1.20 (0.32, 4.48); 0.783 |
| Mild episodes only | 0.96 (0.70, 1.30); 0.770 | 0.96 (0.71, 1.30); 0.788 |
| Mild-to-moderate episodes only | 0.74 (0.45, 1.22); 0.241 | 0.74 (0.45, 1.23); 0.246 |
| **Adjusted for site and enrolment week** | | |
| All episodes | 0.92 (0.71, 1.18); 0.516 | 0.92 (0.71, 1.18); 0.511 |
| Participant-reported episodes only | 0.91 (0.70, 1.18); 0.475 | 0.91 (0.70, 1.18); 0.461 |
| **Adjusted for age and sex** | | |
| All episodes | 0.90 (0.70, 1.16); 0.407 | 0.90 (0.70, 1.16); 0.418 |
| Participant-reported episodes only | 0.89 (0.68, 1.16); 0.386 | 0.89 (0.68, 1.16); 0.389 |
| **Multivariable (backward selection)[3]** | | |
| All Episodes | 0.86 (0.66, 1.11); 0.238 | 0.86 (0.67, 1.11); 0.261 |
| Participant-reported episodes only | 0.85 (0.65, 1.10); 0.218 | 0.85 (0.65, 1.11); 0.235 |

Abbreviations: HR=hazards ratio; 95% CI= 95% confidence interval.

1. Participant with no evidence of an infection episode who completed less than 80% of the expected diary app entries were excluded because we could not be sure that they never had an infection. Participants with inconclusive episodes (N=36) were also removed. Since the outcome is time to first infection, infection episodes that could not be dated dropped out of the model.
2. All participants were included regardless of diary app completion, except participants with inconclusive episodes (N=36). The sensitivity analysis assumed that participant with no evidence of an infection episode who completed less than 80% of the expected diary app entries did in fact never have an infection. Since the outcome is time to first infection, infection episodes that could not be dated dropped out of the model. An additional N=15 participants were dropped with no data regarding follow-up time.
3. Covariates considered as potential confounders are shown in Table S4. Covariates retained in the model were: age in years, additional number of household members, function, % work hours with patient contact, hospital department, expected to work in COVID-ward, past history of BCG vaccination, current use of hypertension medication.

### Table S7: Sensitivity analyses assuming inconclusive infections were or were not true infections

#### A) Logistic regression (cumulative incidence)

| BCG vs placebo | | Analysis population[1]<br>N=1,309<br>298 episodes | Inconclusive as no infection[2]<br>N=1,344<br>298 episodes | Inconclusive as infection[3]<br>N=1,344<br>333 episodes |
|---|---|---|---|---|
| | | **OR (95% CI); p** | **OR (95% CI); p** | **OR (95% CI); p** |
| **Unadjusted** | | | | |
| All episodes | | 0.93 (0.72, 1.20); 0.563 | 0.92 (0.71, 1.19); 0.540 | 0.95 (0.74, 1.21); 0.661 |
| Participant-reported episodes only | | 0.89 (0.66, 1.18); 0.413 | 0.88 (0.66, 1.18); 0.396 | 0.92 (0.70, 1.20); 0.527 |
| Asymptomatic episodes only | | 1.15 (0.62, 2.16); 0.654 | 1.15 (0.62, 2.16); 0.659 | 1.15 (0.62, 2.16); 0.654 |
| Mild episodes only | | 0.94 (0.68, 1.31); 0.714 | 0.93 (0.67, 1.30); 0.677 | 0.94 (0.68, 1.31); 0.714 |
| Mild-to-moderate episodes only | | 0.73 (0.43, 1.23); 0.241 | 0.73 (0.43, 1.22); 0.229 | 0.73 (0.44, 1.23); 0.241 |
| **Adjusted for site, enrolment week** | | | | |
| All episodes | | 0.93 (0.72, 1.20); 0.577 | 0.93 (0.72, 1.20); 0.540 | 0.95 (0.74, 1.21); 0.676 |
| Participant-reported episodes only | | 0.88 (0.66, 1.18); 0.395 | 0.89 (0.66, 1.18); 0.398 | 0.91 (0.69, 1.20); 0.521 |
| **Adjusted for age and sex** | | | | |
| All episodes | | 0.90 (0.70, 1.17); 0.443 | 0.90 (0.69, 1.16); 0.411 | 0.93 (0.72, 1.19); 0.559 |
| Participant-reported episodes only | | 0.86 (0.64, 1.15); 0.304 | 0.85 (0.64, 1.14); 0.283 | 0.90 (0.68, 1.18); 0.429 |
| **Multivariable (backward selection)[4]** | | | | |
| All episodes | | 0.85 (0.65, 1.12); 0.249 | 0.85 (0.65, 1.11); 0.236 | 0.90 (0.70, 1.16); 0.415 |
| Participant-reported episodes only | | 0.81 (0.59, 1.09); 0.161 | 0.80 (0.59, 1.09); 0.155 | 0.87 (0.65, 1.15); 0.317 |
| **Multinomial** | | | | |
| Unadjusted All episodes | Asymptomatic | 1.15 (0.62, 2.14); 0.654 | 0.99 (0.54, 1.79); 0.966 | 1.15 (0.62, 2.14); 0.654 |
| | Mild | 0.94 (0.68, 1.31); 0.714 | 0.93 (0.67, 1.30); 0.677 | 0.94 (0.68, 1.31); 0.714 |
| | Mild-to-moderate | 0.73 (0.44, 1.23); 0.241 | 0.73 (0.44, 1.22); 0.229 | 0.73 (0.44, 1.23); 0.241 |
| Unadjusted Participant-reported only | Asymptomatic | 0.95 (0.27, 3.31); 0.937 | 0.59 (0.19, 1.82); 0.358 | 0.95 (0.27, 3.31); 0.937 |
| | Mild | 0.94 (0.67, 1.32); 0.720 | 0.93 (0.66, 1.31); 0.685 | 0.94 (0.67, 1.32); 0.720 |
| | Mild-to-moderate | 0.73 (0.43, 1.23); 0.236 | 0.72 (0.43, 1.22); 0.224 | 0.73 (0.43, 1.23); 0.236 |
| Adjusted for site, enrolment week All episodes | Asymptomatic | 1.15 (0.62, 2.14); 0.663 | 0.98 (0.54, 1.79); 0.957 | 1.15 (0.62, 2.14); 0.659 |
| | Mild | 0.95 (0.68, 1.32); 0.747 | 0.94 (0.68, 1.31); 0.726 | 0.95 (0.68, 1.32); 0.761 |
| | Mild-to-moderate | 0.73 (0.44, 1.23); 0.243 | 0.73 (0.43, 1.23); 0.237 | 0.74 (0.44, 1.24); 0.249 |
| Adjusted for age and sex All episodes | Asymptomatic | 1.13 (0.61, 2.10); 0.699 | 0.97 (0.53, 1.76); 0.913 | 1.13 (0.61, 2.11); 0.702 |
| | Mild | 0.91 (0.65, 1.27); 0.573 | 0.90 (0.65, 1.25); 0.533 | 0.91 (0.65, 1.27); 0.570 |
| | Mild-to-moderate | 0.73 (0.43, 1.22); 0.230 | 0.72 (0.43, 1.21); 0.212 | 0.73 (0.43, 1.22); 0.227 |
| Multivariable (backward selection)[5] | Asymptomatic | 1.13 (0.60, 2.11); 0.711 | 0.97 (0.53, 1.78); 0.932 | 1.13 (0.60, 2.11); 0.705 |
| | Mild | 0.87 (0.62, 1.22); 0.417 | 0.86 (0.61, 1.21); 0.384 | 0.87 (0.62, 1.22); 0.419 |
| | Mild-to-moderate | 0.71(0.41, 1.21); 0.202 | 0.70 (0.42, 1.19); 0.193 | 0.71 (0.42, 1.21); 0.205 |

Abbreviations: OR=odds ratio; 95% CI= 95% confidence interval.

1. Participants with no evidence of an infection episode who completed less than 80% of the expected diary app entries were excluded because we could not be sure that they never had an infection. Participants with inconclusive episodes (N=36) were also removed.
2. In the randomised population, 36 episodes were coded as inconclusive, and 35 of these were in participants who completed at least 80% of expected diary app entries. These 35 were added to the analysis population as not having had an infection.
3. In the randomised population, 36 episodes were coded as inconclusive, and 35 of these were in participants who completed at least 80% of expected diary app entries. These 35 were added to the analysis population as having had an infection, with unknown severity.
4. Covariates considered as potential confounders are shown in Table S4. Covariates retained in the model were: age in years, additional number of household members, function, % work hours with patient contact, hospital department, expected to work in COVID-ward, past history of BCG vaccination, current use of hypertension medication.
5. Covariates considered as potential confounders are shown in Table S4. Covariates retained in the model were: age in years, hospital function and department, expected to work in COVID-ward, history of BCG vaccination, and current use of hypertension medication.

### B) Cox regression models (time to first infection)

| BCG vs placebo | Analysis population[1]<br><br>N=1,252<br>241 episodes | Inconclusive as no infection[2]<br>N=1,287<br>241 episodes | Inconclusive as infection[3]<br>N=1,252<br>241 episodes |
|---|---|---|---|
| | **HR (95% CI); p** | **HR (95% CI); p-value** | **HR (95% CI); p** |
| **Unadjusted** | | | |
| All episodes | 0.92 (0.72, 1.19); 0.521 | 0.92 (0.71, 1.18); 0.499 | 0.92 (0.72, 1.19); 0.521 |
| Participant-reported episodes only | 0.91 (0.70, 1.19); 0.493 | 0.91 (0.70, 1.18); 0.473 | 0.91 (0.70, 1.19); 0.493 |
| Asymptomatic episodes only | 1.19 (0.32, 4.44); 0.793 | 1.19 (0.32, 4.43); 0.796 | 1.19 (0.32, 4.44); 0.793 |
| Mild episodes only | 0.96 (0.70, 1.30); 0.770 | 0.95 (0.70, 1.29); 0.731 | 0.96 (0.70, 1.30); 0.770 |
| Mild-to-moderate episodes only | 0.74 (0.45, 1.22); 0.241 | 0.73 (0.44, 1.21); 0.228 | 0.74 (0.45, 1.22); 0.241 |
| **Adjusted for site, enrolment week** | | | |
| All episodes | 0.92 (0.71, 1.18); 0.516 | 0.92 (0.71, 1.18); 0.510 | 0.92 (0.71, 1.18); 0.516 |
| Participant-reported episodes only | 0.91 (0.70, 1.18); 0.475 | 0.91 (0.70, 1.18); 0.477 | 0.91 (0.70, 1.18); 0.475 |
| **Adjusted for age and sex** | | | |
| All episodes | 0.90 (0.70, 1.16); 0.407 | 0.89 (0.69, 1.15); 0.381 | 0.90 (0.70, 1.16); 0.407 |
| Participant-reported episodes only | 0.89 (0.68, 1.16); 0.386 | 0.89 (0.68, 1.15); 0.363 | 0.89 (0.68, 1.16); 0.386 |
| **Multivariable[4]** | | | |
| All Episodes | 0.86 (0.66, 1.11); 0.238 | 0.85 (0.66, 1.10); 0.228 | 0.86 (0.66, 1.11); 0.238 |
| Participant-reported episodes only | 0.85 (0.65, 1.10); 0.218 | 0.84 (0.65, 1.10); 0.210 | 0.85 (0.65, 1.10); 0.218 |

Abbreviations: HR=hazards ratio; 95% CI= 95% confidence interval.

1. Participants with no evidence of an infection episode who completed less than 80% of the expected diary app entries were excluded because we could not be sure that they never had an infection. Participants with inconclusive episodes (N=36) were also removed. Since the outcome is time to first infection, infection episodes that could not be dated dropped out of the model.
2. In the randomised population, 36 episodes were coded as inconclusive, and 35 of these were in participants who completed at least 80% of expected diary app entries. These 35 were added to the analysis population as not having had an infection.
3. In the randomised population, 36 episodes were coded as inconclusive, and 35 of these were in participants who completed at least 80% of expected diary app entries. These 35 were added to the analysis population as having had an infection. However, they dropped out of the analysis because none of these infections could be dated.
4. Covariates considered as potential confounders are shown in Table S4. Covariates retained in the model were: age in years, additional number of household members, function, % work hours with patient contact, hospital department, expected to work in COVID-ward, past history of BCG vaccination, current use of hypertension medication.
